# Sphincterotomy for Biliary Sphincter of Oddi Disorder and idiopathic Acute Recurrent Pancreatitis: THE RESPOND LONGITUDINAL COHORT

**DOI:** 10.1101/2024.04.18.24305985

**Authors:** Gregory A. Coté, B. Joseph Elmunzer, Haley Nitchie, Richard S. Kwon, Field F. Willingham, Sachin Wani, Vladimir Kushnir, Amitabh Chak, Vikesh Singh, Georgios Papachristou, Adam Slivka, Martin Freeman, Srinivas Gaddam, Priya Jamidar, Paul Tarnasky, Shyam Varadarajulu, Lydia D. Foster, Peter B. Cotton

**Affiliations:** Department of Medicine, Division of Gastroenterology & Hepatology, Oregon Health & Science University; Department of Medicine, Division of Gastroenterology & Hepatology, Medical University of South Carolina, Charleston, USA; Department of Internal Medicine, Division of Gastroenterology & Hepatology, University of Michigan, Ann Arbor, MI; Department of Medicine, Division of Digestive Diseases, School of Medicine, Emory University, Atlanta, GA; Department of Medicine, Division of Gastroenterology and Hepatology, University of Colorado Anschutz Medical Campus, Aurora, Colorado, USA; Department of Medicine, Division of Gastroenterology, Washington University in St. Louis School of Medicine, St. Louis, MO; Digestive Health Institute, Department of Medicine, Case Western Reserve University, Cleveland, OH; Division of Gastroenterology, Johns Hopkins Medical Institutions, Baltimore, MD; Department of Medicine, Division of Gastroenterology & Hepatology, Ohio State University, Columbus, OH; Department of Medicine, University of Pittsburgh, Pittsburgh, PA; Department of Medicine, University of Minnesota, Minneapolis, MN; Department of Medicine, Cedars Sinai School of Medicine; Department of Medicine, Yale University School of Medicine, New Haven, CT; Methodist Digestive Institute, Dallas, TX; Digestive Health Institute, Orlando Health, Orlando, FL; Department of Public Health Science, Medical University of South Carolina, Charleston, USA

**Keywords:** Sphincter of Oddi Dysfunction, Cholangiopancreatography, Endoscopic Retrograde, abdominal pain, acute pancreatitis, Quality of Life, Sphincterotomy

## Abstract

**Objective:** Sphincter of Oddi Disorders (SOD) are contentious conditions in patients whose abdominal pain, idiopathic acute pancreatitis (iAP) might arise from pressurization at the sphincter of Oddi. The present study aimed to measure the benefit of sphincterotomy for suspected SOD.

**Design:** Prospective cohort conducted at 14 U.S. centers with 12 months follow-up. Patients undergoing first-time ERCP with sphincterotomy for suspected SOD were eligible: pancreatobiliary-type pain with or without iAP. The primary outcome was defined as the composite of improvement by Patient Global Impression of Change (PGIC), no new or increased opioids, and no repeat intervention. Missing data were addressed by hierarchal, multiple imputation scheme.

**Results:** Of 316 screened, 213 were enrolled with 190 (89.2%) of these having a dilated bile duct, abnormal labs, iAP, or some combination. By imputation, an average of 122/213 (57.4% [95%CI 50.4-64.4]) improved; response rate was similar for those with complete follow-up (99/161, 61.5%, [54.0-69.0]); of these, 118 (73.3%) improved by PGIC alone. Duct size, elevated labs, and patient characteristics were not associated with response. AP occurred in 37/213 (17.4%) at a median of 6 months post-ERCP and was more likely in those with a history of AP (30.9 vs. 2.9%, p<0.0001).

**Conclusion:** Nearly 60% of patients undergoing ERCP for suspected SOD improve, although the contribution of a placebo response is unknown. Contrary to prevailing belief, duct size and labs are poor response predictors. AP recurrence was common and like observations from prior non-intervention cohorts, suggesting no benefit of sphincterotomy in mitigating future AP episodes.

Key Messages

WHAT IS ALREADY KNOWN ON THIS TOPIC

- It is not clear if the sphincter of Oddi can cause abdominal pain (Functional Biliary Sphincter of Oddi Disorder) and idiopathic acute pancreatitis (Functional Pancreatic Sphincter of Oddi Disorder), and whether ERCP with sphincterotomy can ameliorate abdominal pain or pancreatitis.

WHAT THIS STUDY ADDS

- Using multiple patient-reported outcome measures, most patients with suspected sphincter of Oddi disorder improve after ERCP with sphincterotomy.
- Duct size, elevated pancreatobiliary labs, and baseline patient characteristics are not independently associated with response.
- There is a high rate of recurrent acute pancreatitis within 12 months of sphincterotomy in those with a history of idiopathic acute pancreatitis.

HOW THIS STUDY MIGHT AFFECT RESEARCH, PRACTICE, OR POLICY

- Since a discrete population with a high (> 80-90%) response rate to sphincterotomy for suspected pancreatobiliary pain could not be identified, there is a need for additional observational and interventional studies that include phenotyping of patients using novel imaging or biochemical biomarkers.
- There remains a pressing need for quantitative nociceptive biomarkers to distinguish pancreatobiliary pain from other causes of abdominal pain or central sensitization.
- Discovery of blood-, bile-, or imaging-based biomarkers for occult microlithiasis and pancreatitis may be helpful in predicting who is likely to benefit from sphincterotomy.

## Introduction

Abdominal pain is common and often associated with overlapping disorders of gut-brain interaction (DGBI). Functional sphincter of Oddi disorders (SOD) are defined by episodic abdominal pain that bear a resemblance to discomfort originating from the biliary tree, pancreas, or both,^1^ but equating pain unequivocally with SOD is not possible.^2^ Few disorders incite more controversy than SOD since there is no diagnostic test to prove that the sphincter is the source of pain and prospective studies evaluating the benefit of endoscopic sphincterotomy have been inconsistent.^3–6^

Physician-defined characteristics of SOD include the presence of a dilated common bile duct (CBD), elevated liver or pancreas chemistries associated with pain,^7^ and a more discrete subtype which are those idiopathic recurrent acute pancreatitis (iRAP). The latter represents a diagnostic spectrum ranging from radiographic changes definitive for acute pancreatitis to only pain with elevated pancreas biochemistries.^8^ In fact, experts do not consider pain associated with a dilated duct and abnormal biochemistries as SOD: this has been renamed papillary stenosis and sphincterotomy is recommended based on historical studies.^5^

Physician-defined characteristics of SOD are not as objective as originally conceived. Duct dilation is an imperfect clinical biomarker given the prevalence of physiologic dilation that occurs with cholecystectomy, aging, and opioid use.^9^ Similarly, the specificity of elevated liver chemistries as an SOD indicator is compromised by the high baseline rate of metabolic dysfunction associated steatotic liver disease (MASLD, estimated prevalence 10-35%)^10^ as well as abnormal aminotransferase levels in the general population (estimated prevalence 10-30%).^11–13^ Finally, iRAP is a subjective diagnosis since physicians commonly diagnose acute pancreatitis in the absence of radiographic changes and deeming a case idiopathic varies across clinical practices and individual cases.

Given inconsistent outcomes and variability in current clinical practice, the Results of ERCP for SPhincter of Oddi Disorders (RESPOnD) was a multicenter, prospective cohort study designed to measure the 12-month response to sphincterotomy, as defined by patient-reported outcome measures (PROMs), among patients undergoing ERCP for suspected SOD.

## Methods

### Study Design, Setting, and Participants

RESPOnD was a prospective cohort study conducted at 14 U.S. medical centers. Eligible patients included those age ≥ 18 years and offered ERCP with sphincterotomy for suspected SOD and no history of endoscopic sphincterotomy between January 2018 and March 2022. This included all SOD subtypes: 1) Biliary: pain with a dilated bile duct, elevated liver chemistries, or both, 2) Pancreatic: idiopathic acute pancreatitis, diagnosed radiographically or by elevated pancreas chemistries alone, and 3) Mixed type: characteristics of both Biliary and Pancreatic. For centers participating in the Stent vs. Indomethacin trial (SVI),^14^ patients could be enrolled after their ERCP and then followed prospectively to the primary endpoint. Patients with chronic pancreatitis, pregnancy, or a psychiatric disorder that precluded obtaining informed consent were excluded ***(study protocol is available in supplemental materials)***. A history of cholecystectomy was not required for participation if the treating physician recommended ERCP as the initial intervention. Similarly, sphincter of Oddi manometry was not required since most participating providers performed empiric sphincterotomies and manometry’s poor reproducibility and clinical utility.^15^ The study was reviewed and approved by the Human Subjects Protection Office at each participating center before commencement of enrollment.

After signing informed consent, patients completed a baseline assessment before the ERCP procedure during which they completed a series of validated questionnaires designed to characterize their pain, pain-related disability, relevant medical history, and factors such as underlying somatization, depression, anxiety, expectation of response, and recent opioid utilization ***(supplementary table 1)***. Bile duct diameter was recorded using magnetic resonance cholangiopancreatography, computed tomography, and ERCP, with dilation being defined as ≥ 12mm on any.^6^ A less stringent definition for bile duct dilation was considered but did not impact the results significantly (data not shown). Similarly, an abnormal biochemistry was defined by a liver biochemistry (AST, ALT, or alkaline phosphatase) > 2x upper limit of normal (ULN) or > 3x ULN for pancreas chemistry (amylase or lipase).

During ERCP, the treating physician performed biliary, pancreatic, or dual sphincterotomies per their clinical judgement. Patients received periprocedural rectal indomethacin and a prophylactic pancreatic duct stent was placed per clinical judgement or per treatment allocation in the SVI trial if applicable.

Following ERCP, patients were followed quarterly in-person or by telephone for 12 months. During these visits, patients completed a series of validated, patient-reported outcome measures, queried for recent opioid and other medication use for their abdominal symptoms, hospitalizations for abdominal symptoms, and whether additional interventions were performed for their symptoms.^16^

### Primary and secondary outcomes

The primary outcome definition of success was determined at 12 months and was a composite of the Patient Global Impression of Change (PGIC) rated as “much improved” or “very much improved”, without the need for a repeat ERCP or other abdominal intervention, and the same or less days of prescription opioid use during month 12 compared to the month preceding the index ERCP. Secondary outcomes included the change (from baseline) in pain-related disability measured by the Recurrent Abdominal Pain Intensity and Disability (RAPID) instrument,^17^ physical and mental health summary scores as well as sub-scores derived from PROMIS-29.^18^ Acute pancreatitis, defined by revised Atlanta criteria^19^, was deemed non-iatrogenic unless it occurred within 30 days of the index or a follow-up ERCP.

### Statistical analysis

The original sample size estimate was 360 subjects enrolled at sites participating in the SVI trial. This was based on SVI’s original sample size and rate of treatment naïve subjects enrolled in SVI at the time of this study’s inception (33%); RESPOnD enrollment began 28 months after SVI. A sample size of 360 would have resulted in a confidence interval width on our estimated proportion of success to be approximately 10±5%. Enrollment in RESPOnD was lower than anticipated because of a smaller proportion of eligible subjects in SVI (24% after commencement of RESPOnD) and the COVID-19 pandemic. Recruitment was terminated early due to funding expiration and analysis demonstrating a confidence interval width like our original estimate.

The prespecified primary analysis was an unadjusted primary outcome success proportion with a two-sided 95% confidence interval. Secondary outcomes were assessed for association with the primary outcome via a generalized linear model with log link and Poisson distribution, with the primary outcome as the dependent variable and the secondary outcome as the independent variable. Models were adjusted for the baseline value of the secondary outcome. The Poisson distribution was used due to convergence issues with the prespecified binomial distribution. For safety outcomes, the unadjusted proportion and two-sided 95% confidence interval was estimated.

For subjects missing primary outcome data, we followed a hierarchical imputation scheme, which included multiple imputation. Multiple imputation assumed data were missing at random and a sensitivity analysis was conducted to assess the impact of missing data; multiple imputation allows for the uncertainty of missing data by creating several different plausible imputed data sets and appropriately combining results obtained from each of them.^20^

Two multivariate logistic models associated with the imputed primary outcome were constructed. The first model considered physician-defined characteristics of SOD: elevated liver chemistry >2x ULN, bile duct diameter ≥ 12mm, and history of idiopathic acute pancreatitis. The second model considered baseline patient characteristics suspected to have an association with the primary outcome by univariate testing (p ≤ 0.10).

## Results

### Enrollment and follow-up

During the study period, 316 patients were screened for eligibility and 216 were consented to participate. Three were excluded due to inability to complete ERCP (2) or missing informed consent documentation (1). Of the remaining 213 subjects, 179 (84.0%) were enrolled prior to their index ERCP and 34 (16.0%) after ERCP but prior to their assessment of the primary outcome **(*figure 1*)**. The baseline demographics of these two subgroups were similar (data not shown). A total of 169 (79.3%) completed 12-month follow-up, 36 (16.9%) were lost to follow-up, 5 (2.3%) withdrew consent, and 3 (1.4%) died.

**Figure 1.**
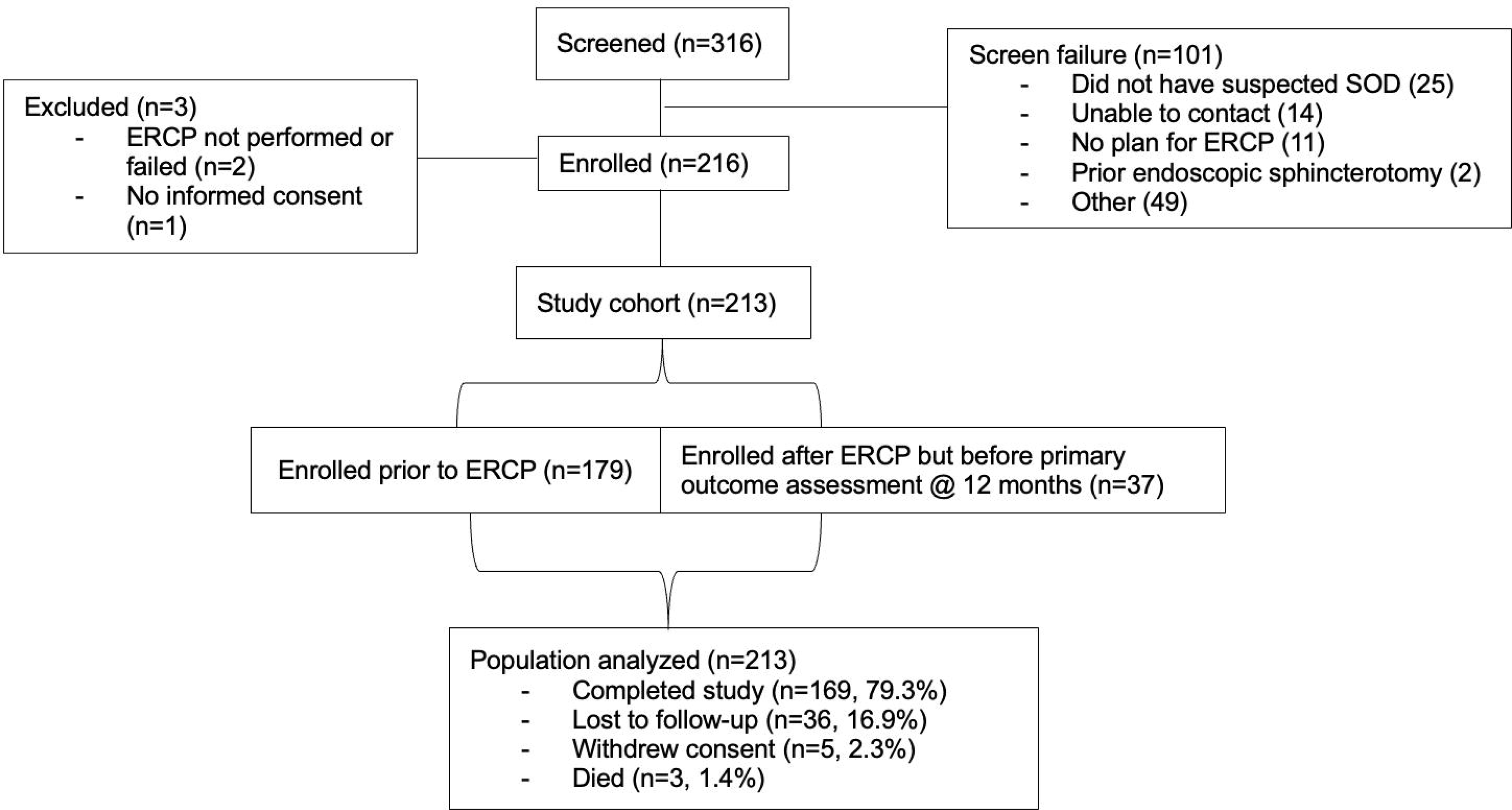
Screening and enrollment. Summary of enrollment and retention in RESPOnD. Some patients (n=37) were enrolled after their ERCP but before the 12-month primary outcome assessment. In these cases, the baseline assessment did not include completion of validated instruments characterizing their pain or psychosocial comorbidity. These subjects completed all other baseline and follow-up assessments per protocol.

### Baseline characteristics

The cohort was predominantly female sex (80.8%), non-Hispanic White (82.6%), and middle aged (50.6 ± 14.9 years, ***table 1***). 110/213 (51.6%) had a history of at least one episode of acute pancreatitis in their lifetime, 99 (46.5%) within 12 months of their ERCP. The majority had previously undergone a cholecystectomy, were nonsmokers at the time of enrollment, and drank little to no alcohol. A substantial number of patients had concomitant comorbidities suspected to confound their clinical presentation with abdominal pain, abnormal liver chemistries, or both, including irritable bowel syndrome (41.8%), MASLD (19.7%), fibromyalgia (14.1%), and gastroparesis (8.0%). Most (190/213, 89.2%) subjects presented with one or more physician-defined characteristics of SOD (duct dilation, abnormal pancreatobiliary biochemistries, and/or acute pancreatitis history), leaving 23 (10.8%) with none of these features, suggestive of type III SOD using the Rome III definition.

**Table 1.**
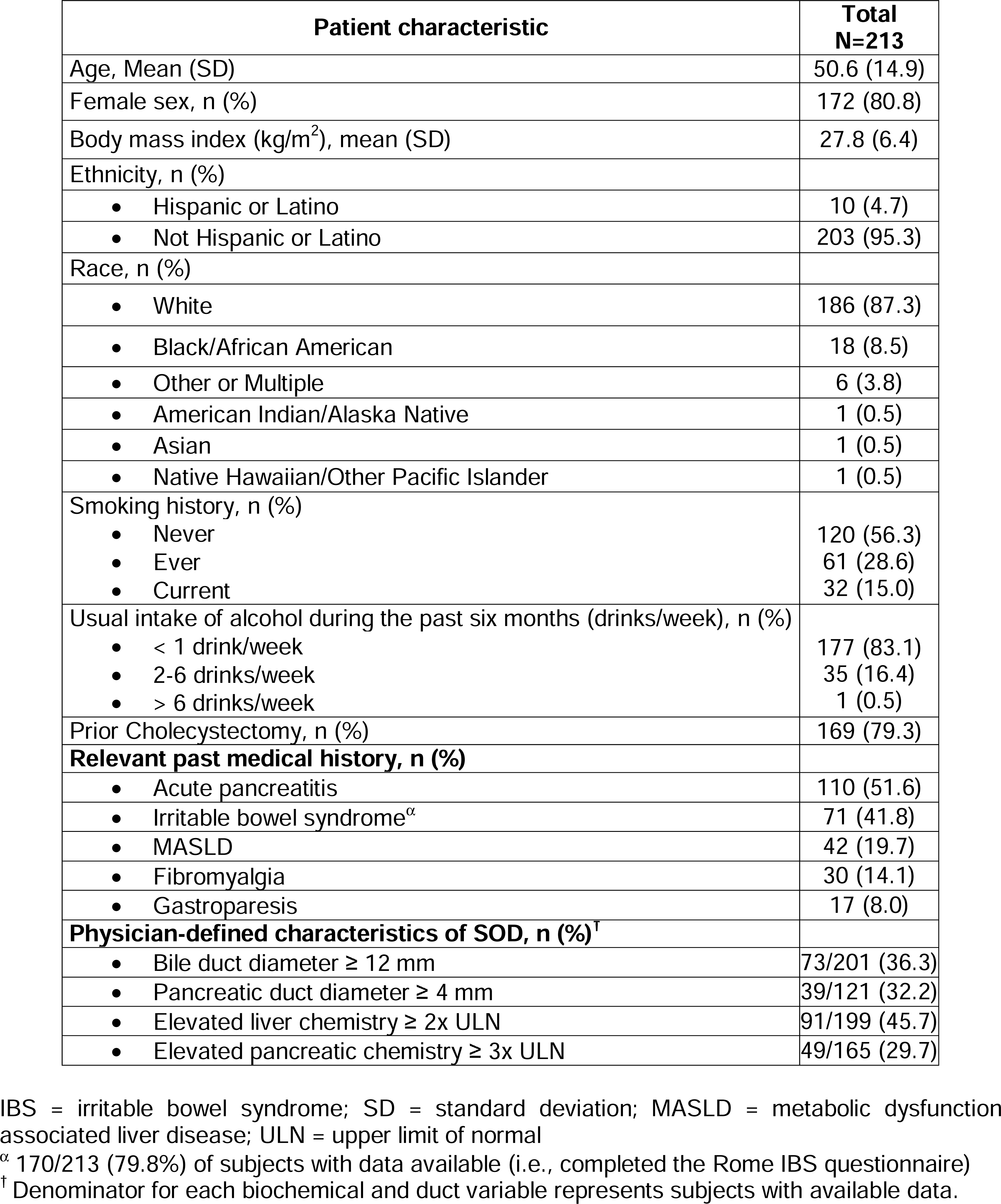
Patient characteristics.

| **Patient characteristic** | **Total**<br>**N=213** |
| --- | --- |
| Age, Mean (SD) | 50.6 (14.9) |
| Female sex, n (%) | 172 (80.8) |
| Body mass index (kg/m<sup>2</sup>), mean (SD) | 27.8 (6.4) |
| Ethnicity, n (%) |  |
| • Hispanic or Latino | 10 (4.7) |
| • Not Hispanic or Latino | 203 (95.3) |
| Race, n (%) |  |
| • White | 186 (87.3) |
| • Black/African American | 18 (8.5) |
| • Other or Multiple | 6 (3.8) |
| • American Indian/Alaska Native | 1 (0.5) |
| • Asian | 1 (0.5) |
| • Native Hawaiian/Other Pacific Islander | 1 (0.5) |
| Smoking history, n (%) |  |
| • Never | 120 (56.3) |
| • Ever | 61 (28.6) |
| • Current | 32 (15.0) |
| Usual intake of alcohol during the past six months (drinks/week), n (%) |  |
| • < 1 drink/week | 177 (83.1) |
| • 2-6 drinks/week | 35 (16.4) |
| • > 6 drinks/week | 1 (0.5) |
| Prior Cholecystectomy, n (%) | 169 (79.3) |
| **Relevant past medical history, n (%)** |  |
| • Acute pancreatitis | 110 (51.6) |
| • Irritable bowel syndrome<sup>α</sup> | 71 (41.8) |
| • MASLD | 42 (19.7) |
| • Fibromyalgia | 30 (14.1) |
| • Gastroparesis | 17 (8.0) |
| **Physician-defined characteristics of SOD, n (%)<sup>†</sup>** |  |
| • Bile duct diameter ≥ 12 mm | 73/201 (36.3) |
| • Pancreatic duct diameter ≥ 4 mm | 39/121 (32.2) |
| • Elevated liver chemistry ≥ 2x ULN | 91/199 (45.7) |
| • Elevated pancreatic chemistry ≥ 3x ULN | 49/165 (29.7) |

IBS = irritable bowel syndrome; SD = standard deviation; MASLD = metabolic dysfunction associated liver disease; ULN = upper limit of normal
<sup>α</sup> 170/213 (79.8%) of subjects with data available (i.e., completed the Rome IBS questionnaire)
<sup>†</sup> Denominator for each biochemical and duct variable represents subjects with available data.

The majority (143/169, 84.6%) had some degree of abdominal pain within 7 days of their index ERCP (***table 2***); the median number of pain days was 30 in the last 90, rated as a median of 7/10 and resulting in a median pain burden of 180 (interquartile range: 60, 495). Most patients characterized their pain as nociceptive (112/168, 66.7%). Approximately 1/3^rd^ of patients had low physical or mental health, whereas nearly half of participants had a high likelihood of somatization. Opioids were used within 30 days of ERCP in 62/171 (36.3%). The majority (90/166, 54.2%) expected their pain to be very much better or completely gone to consider the procedure as successful.

**Table 2.** Pre-procedure pain characteristics, analgesic use, and psychiatric comorbidities.

| Characteristic | Denominator |  |
| --- | --- | --- |
| **Pain characteristic** |  |  |
| Response to cholecystectomy, n (%) |  |  |
| • Complete response | 161 | 67/161 (41.6) |
| • Partial response |  | 36/161 (22.4) |
| • No response |  | 58/161 (36.0) |
| Current pain like before cholecystectomy, n (%) | 162 | 94/162 (58.0) |
| • Days of pain episodes in past 90 days, median (IQR) (*n*=169) | 169 | 30 (8, 90) |
| • Any abdominal pain in the last 7 days, n (%) | 169 | 143 (84.6) |
| • Pain intensity (0-10) in past 90 days, median (IQR) | 168 | 7 (6, 9) |
| • Pain burden (frequency x intensity product) in past 90 days, median (IQR) | 168 | 180 (60, 495) |
| • Neuropathic pain, n (%) | 167 | 32 (19.2) |
| • Nociceptive pain, n (%) | 168 | 112 (66.7) |
| **Psychosocial characteristic** |  |  |
| • High likelihood of depression*, n (%) | 167 | 21 (12.6) |
| • High likelihood of anxiety<sup>ψ</sup>, n (%) | 167 | 35 (21.0) |
| • High likelihood of somatization, n (%) | 167 | 76 (45.5) |
| • Low physical health<sup>†</sup>, n (%) | 165 | 60 (36.4) |
| • Low mental health<sup>‡</sup>, n (%) | 165 | 52 (31.5) |
| **Medication use in the 30 days prior to ERCP** |  |  |
| • Opioid, n (%) | 171 | 62 (36.3) |
| • NSAIDs, n (%) | 171 | 74 (43.3) |
| • Antispasmodics, n (%) | 171 | 48 (28.1) |
| • Neuromodulators, n (%) | 171 | 83 (48.5) |

\* High likelihood of depression defined by a BSI-18 Depression T score ≥ 65
<sup>ψ</sup> High likelihood of anxiety defined by BSI-18 Anxiety T score ≥ 65
<sup>†</sup> Low physical health defined as a PROMIS-29 Physical Health Summary T score ≤ 40
<sup>‡</sup> Low mental health defined as a PROMIS-29 Mental Health Summary T score ≤ 4

### Procedure characteristics

During ERCP, only 2/212 (0.9%) of patients underwent sphincter of Oddi manometry. However, most patients underwent a biliary sphincterotomy (204/213, 95.8%), pancreatic sphincterotomy (26/213, 12.2%), or both (20, 9.4%); 2 (0.9%) underwent a precut sphincterotomy alone and 1 (0.5%) did not have any sphincterotomy performed. A serious adverse event developed within 30 days of the index ERCP in 68/213, (31.9%); 22/213 (10.3%) developed post-ERCP pancreatitis and 1/213 (0.5%) perforation ***(supplementary table 2)***.

### Primary outcome: improvement following ERCP

Among 213 patients who underwent ERCP, an average of 122 (57.4% [95%CI 50.4-64.4]) met the imputed primary outcome for success at 12-month follow-up. The response rate changed minimally using the partial hierarchal imputation method: 103/179 (57.5% [50.3-64.8]). In a sensitivity analysis restricted to patients with complete follow-up data (n=161), the response rate was similar (99/161, 61.5%, [54.0-69.0]). Of those with complete follow-up data, 118 (73.3%) met the primary success by PGIC alone but 19 failed due to the need for additional procedures (n=15), new or increased need for opioids (n=3), or both (n=1).

### Secondary outcome: development of acute pancreatitis > 30 *days after ERCP*

During 12-month follow-up, 37/213 (17.4%) developed at least one episode of non-iatrogenic acute pancreatitis at a median of 6 months after the index ERCP. Non-iatrogenic acute pancreatitis was more likely if the patient had a history of acute pancreatitis prior to the index ERCP (30.9%, p<0.0001, ***figure 2***), but some events occurred in those with no prior history of acute pancreatitis (2.9%).

**Figure 2.**
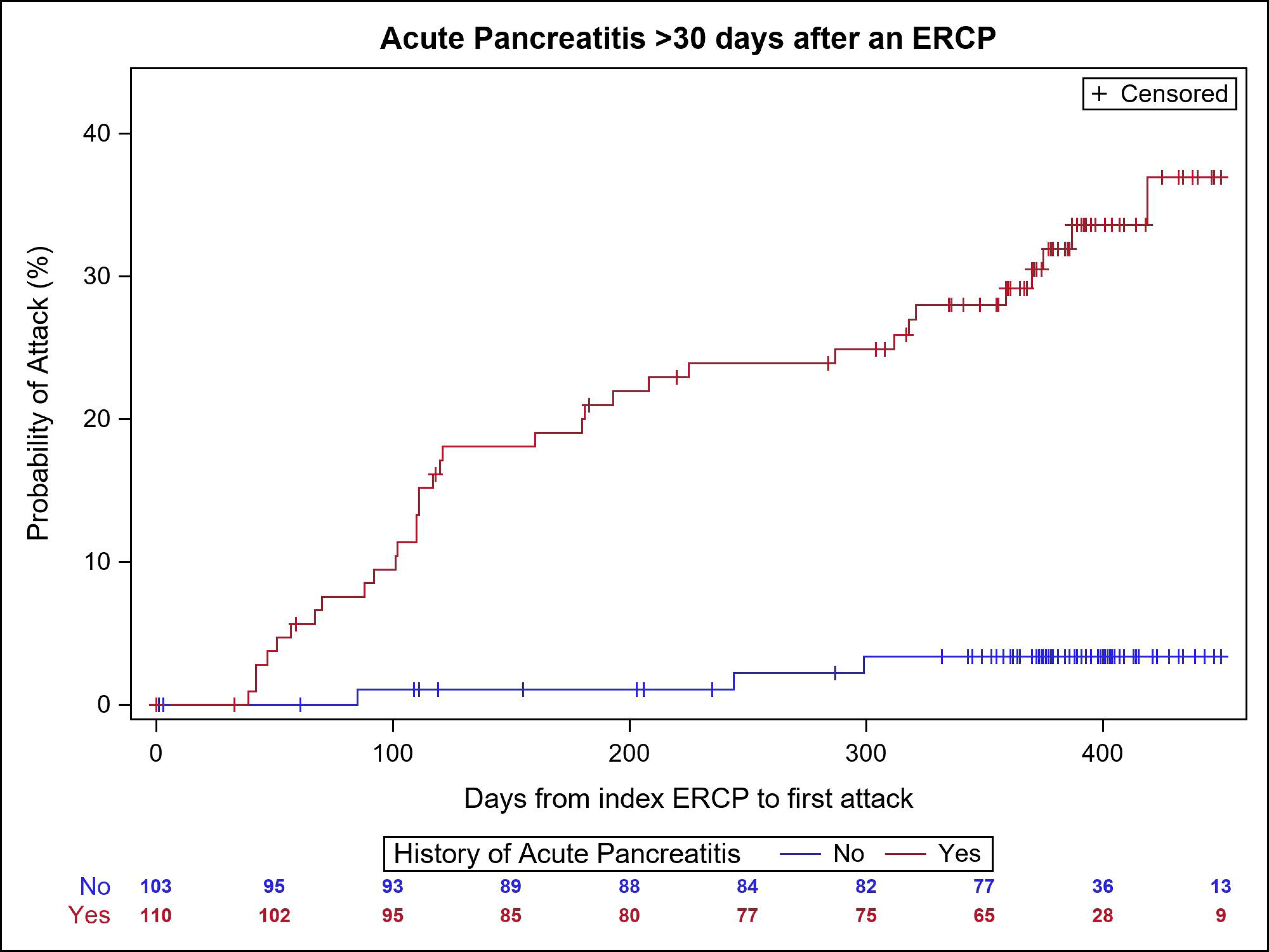
Development of acute pancreatitis following ERCP for suspected SOD. Time to acute pancreatitis after the index ERCP with sphincterotomy. Episodes which occurred within 30 days of the index or follow-up ERCP are not represented.

### Relationship between physician defined SOD, patient characteristics, and outcome

There were no significant differences in success rates when patients were classified using the physician-defined criteria for SOD, with success rates in biliary type I (51.3%), type II (51.6%), type III (53.6%), pancreatic (56.1%), and mixed type (67.5%) being similar (p=0.4152) (***table 3***). Similarly, a history of acute pancreatitis (60.7 vs. 53.8%, p=0.3560), an elevated liver chemistry > 2x ULN (60.4 vs. 56.1%, p=0.5627), and bile duct dilation ≥ 12mm (56.8 vs. 57.6%, p=0.8062) did not correspond with a higher likelihood of success. Multiple patient characteristics potentially associated with response to sphincterotomy were evaluated by univariate analysis, with key comparisons summarized in ***table 4*.** The most significant negative associations with the primary outcome were prescription opioid use in the last 30 days (46.8 vs. 66.3%, p=0.0236), low baseline physical health (50.0 vs. 66.2%, p=0.0512), and high likelihood of somatization at the time of index ERCP (52.6 vs. 67.0%, p=0.0750). There was no association between response to ERCP and a past medical history of irritable bowel syndrome, MASLD, gastroparesis, smoking, and pain subtype (nociceptive or neuropathic).

**Table 3.** Physician defined characteristics of SOD and response to ERCP treatment.

|  | Physician-defined characteristic |  | Denominator | Success, n (%)<sup>Ψ</sup> | p-value |
| --- | --- | --- | --- | --- | --- |
| SOD Subtype | **Biliary SOD alone**<br>• **Elevated liver chemistry > 2x ULN**<br>• **Bile duct diameter ≥ 12 mm** | Both (type I) | 19 | 9.8 (51.3) | 0.4152 |
|  |  | Either (type II) | 54 | 27.9 (51.6) |  |
|  |  | Neither (type III) | 25 | 13.4 (53.6) |  |
|  | **Pancreatic SOD alone** |  | 59 | 33.1 (56.1) |  |
|  | **Mixed type<sup>†</sup>** |  | 55 | 37.1 (67.5) |  |
| **Any history of acute pancreatitis** |  | No | 103 | 55.4 (53.8) | 0.3560 |
|  |  | Yes | 110 | 66.8 (60.7) |  |
| **Elevated liver chemistry > 2x ULN** |  | No | 108 | 60.6 (56.1) | 0.5627 |
|  |  | Yes | 91 | 55.0 (60.4) |  |
| **Bile duct diameter ≥ 12mm** |  | No | 128 | 73.7 (57.6) | 0.8062 |
|  |  | Yes | 73 | 41.5 (56.8) |  |
| SOD = Sphincter of Oddi Disorder; ULN = upper limit of normal |  |  |  |  |  |
| † Patients having characteristics of biliary type I or II and pancreatic SOD (acute pancreatitis history with or without elevated pancreas chemistry > 3x ULN) |  |  |  |  |  |
| Ψ These data represent a composite of 20 imputed datasets, resulting in a reported “n” that is the average of the primary outcome. |  |  |  |  |  |

**Table 4.**
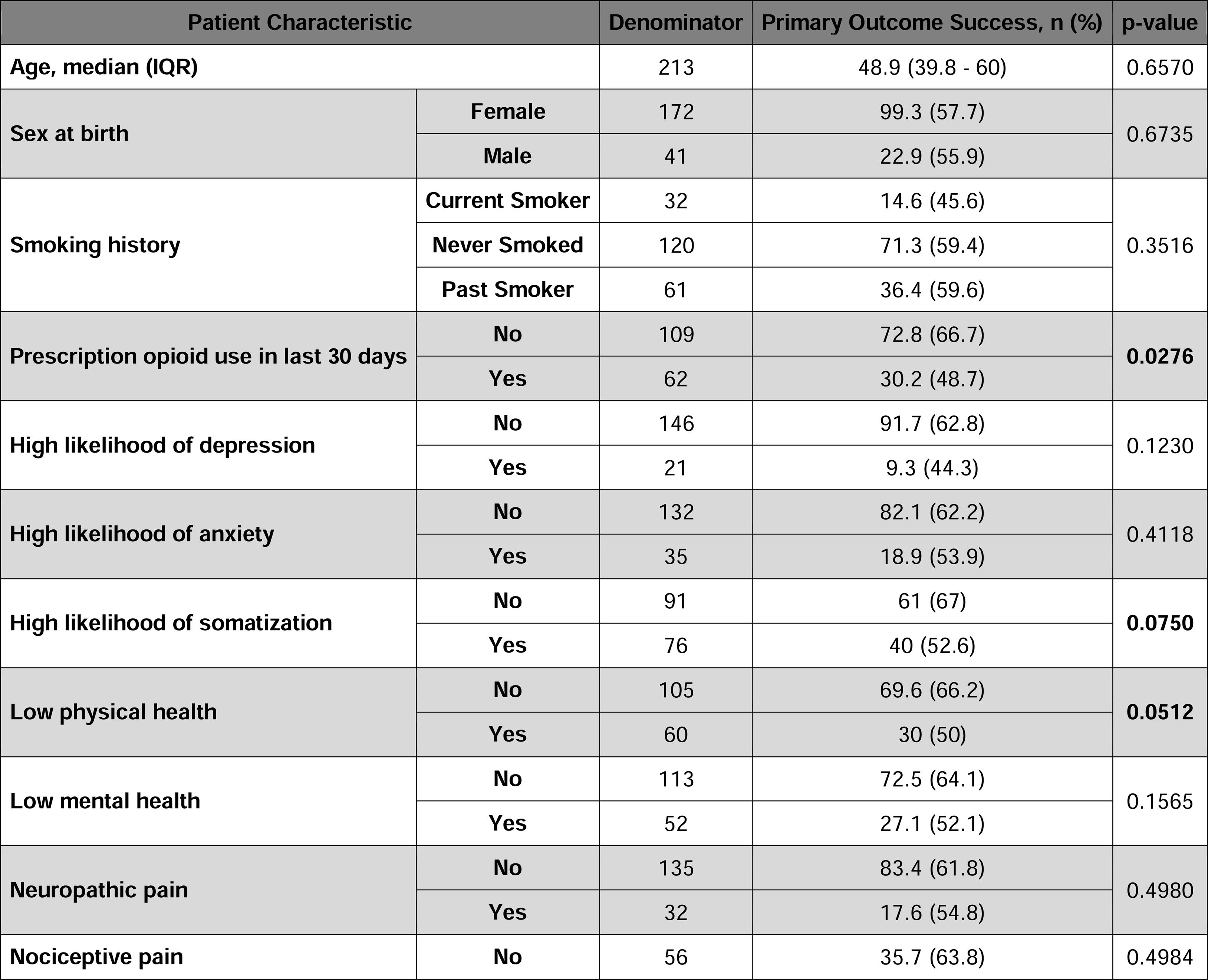
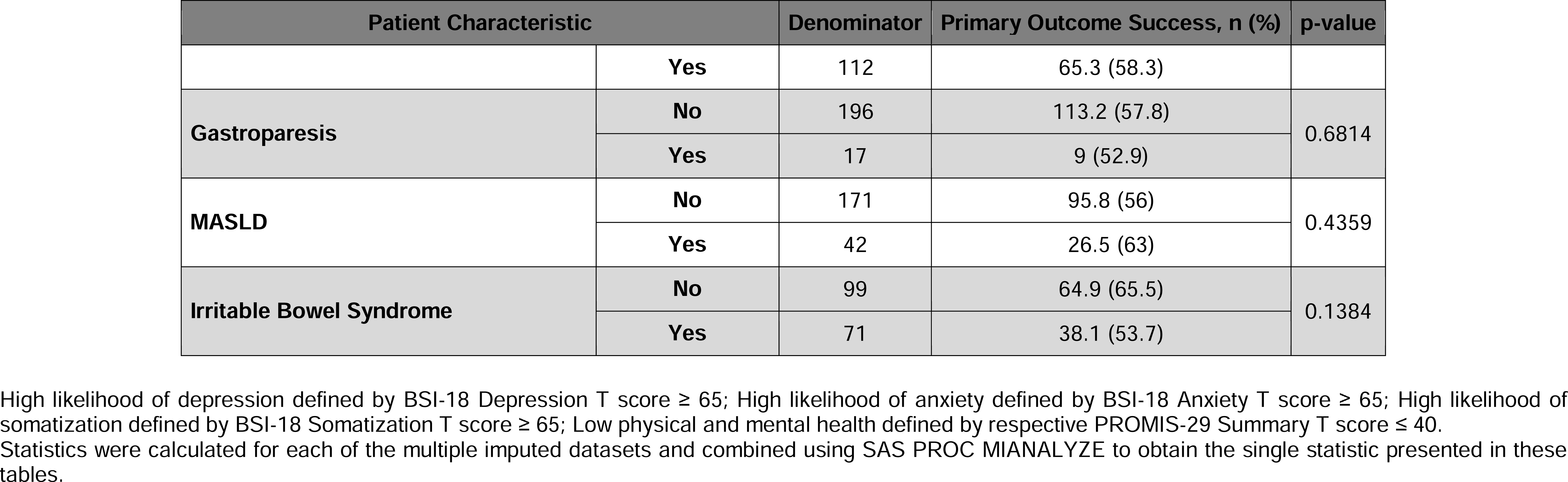
Relationship between patient characteristics and response to ERCP (odds ratio)

### Multivariate analysis

Physician-defined characteristics of SOD were not associated with a higher likelihood of response to ERCP with sphincterotomy ***(table 5)***. In a regression model which considered patient characteristics associated univariately (p ≤ 0.10) with response, none of these factors proved to be independently associated with improvement. Recent opioid use (multivariate odds ratio 0.53 [0.26 – 1.08], p=0.0810) appeared to be the most important patient factor but did not remain statistically significant in the final model.

**Table 5.**
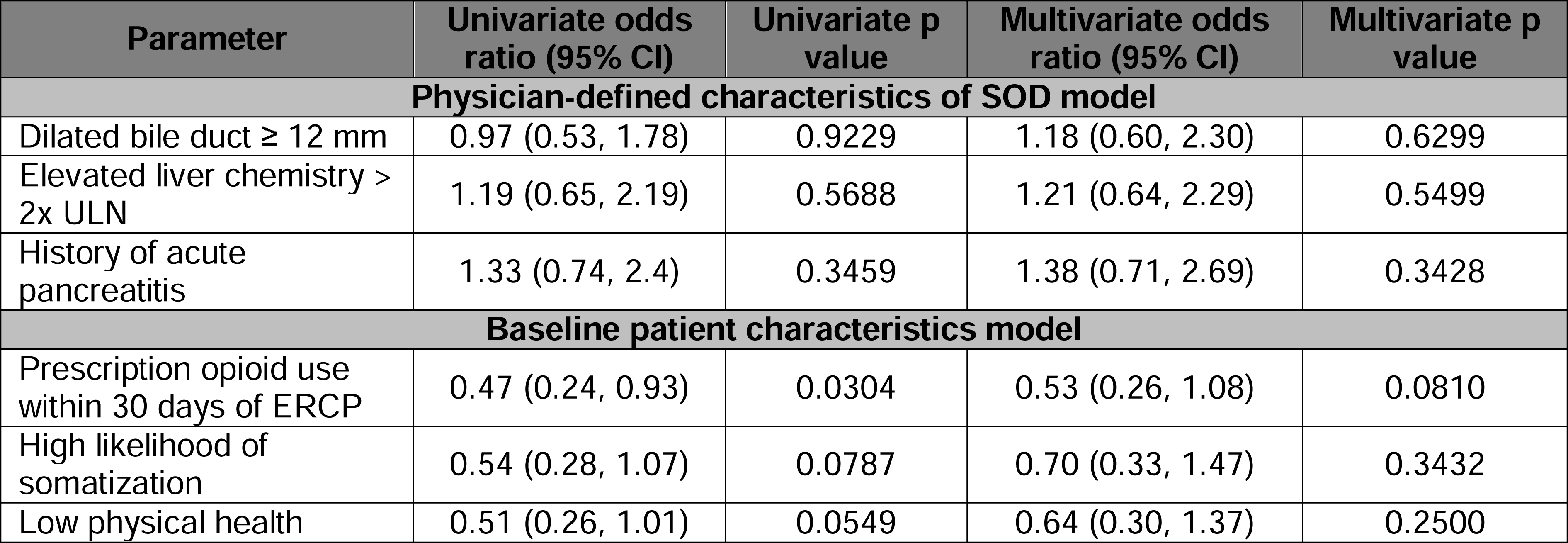
Factors associated with response to ERCP.

| Parameter | Univariate odds ratio (95% CI) | Univariate p value | Multivariate odds ratio (95% CI) | Multivariate p value |
| --- | --- | --- | --- | --- |
| **Physician-defined characteristics of SOD model** |  |  |  |  |
| Dilated bile duct $\geq 12$ mm | 0.97 (0.53, 1.78) | 0.9229 | 1.18 (0.60, 2.30) | 0.6299 |
| Elevated liver chemistry > 2x ULN | 1.19 (0.65, 2.19) | 0.5688 | 1.21 (0.64, 2.29) | 0.5499 |
| History of acute pancreatitis | 1.33 (0.74, 2.4) | 0.3459 | 1.38 (0.71, 2.69) | 0.3428 |
| **Baseline patient characteristics model** |  |  |  |  |
| Prescription opioid use within 30 days of ERCP | 0.47 (0.24, 0.93) | 0.0304 | 0.53 (0.26, 1.08) | 0.0810 |
| High likelihood of somatization | 0.54 (0.28, 1.07) | 0.0787 | 0.70 (0.33, 1.47) | 0.3432 |
| Low physical health | 0.51 (0.26, 1.01) | 0.0549 | 0.64 (0.30, 1.37) | 0.2500 |

### Secondary outcomes

Success defined using the prespecified primary outcome corresponded with change in pain-related disability (mean change in RAPID score from baseline, −41.2 vs. −28.6 for those who failed the primary outcome, p = 0.0127). There was no association between change in RAPID score and bile duct diameter in the complete case population: consistent with the analysis of outcome according to Rome III SOD subtype, there was no correlation between increasing bile duct diameter and reduction in RAPID score ***(p=0.6303, supplemental figure 1).*** Summary physical (6.0 vs. 2.8, p = 0.0294) and mental (7.6 vs. 3.2, p=0.0140) health scores improved significantly among patients meeting the study’s primary outcome for success. In the complete case population and except for anxiety, change in PROMIS-29 sub-scores was significantly associated with the study’s primary outcome ***(supplementary table 3)***. At the 12-month visit, the number of patients who required opioids for abdominal pain in the past 30 days decreased from 62/171 (36.5%) to 24/169 (14.2%).

## Discussion

### Sphincterotomy for suspected pancreatobiliary pain

Is SOD a fabricated etiology for abdominal pain? Many clinicians do not believe SOD is real, and even fewer admit to performing ERCP to ameliorate it (at least by coding data).^21, 22^ The RESPOnD cohort represents a population who share a common characteristic: their treating physician interpreted their abdominal pain and other signs as concerning for SOD and recommended an ERCP with sphincterotomy to address them. This is a common problem, especially in the U.S. where > 1 million cholecystectomies are performed annually and a growing proportion return for a first-time ERCP > 1 year postoperatively; these individuals have higher rates of morbidity following these delayed ERCPs, likely because many have SOD-like characteristics.^23^ Clinicians have the daunting task of trying to determine the etiology of a patient’s abdominal pain when it is clear that pain may manifest similarly yet originate from multiple organs and is often confounded by overlapping disorders of gut brain interaction.^2, 24–26^ This issue of cross-sensitization imposes a substantial challenge in differentiating pain originating from the pancreas, biliary tree, or another organ. A reliable blood or imaging test to confirm that pain is originating in the gallbladder, sphincter of Oddi, or pancreas is lacking, and RESPOnD confirms that standard pancreatobiliary chemistries and duct size are inadequate prognosticators. Last, patients with abdominal pain severe or sustained enough to prompt referral to RESPOnD providers are at-risk for central sensitization;^27^ this results in heightened pain signaling anywhere along the nervous system and probably influences response to local interventions such as sphincterotomy. Despite these challenges, most patients improved after 12 months of follow-up using RESPOnD’s primary outcome definition and numerous secondary outcomes derived from PROMs.

The present study attempts to address the hypothesis that SOD is a cause of abdominal pain by showing that ERCP with sphincterotomy can ameliorate these symptoms. Is the observed response rate – 57% using the pre-defined primary outcome and as high as 73% when using the PGIC alone – too high for a placebo? The placebo response is greatest for patients suffering from the most severe pain and interventions (as opposed to drugs) have a greater placebo effect, especially when offered as pain treatments.^28–31^ In the “Evaluating Predictors & Interventions in Sphincter of Oddi Dysfunction” (EPISOD) study which enrolled patients with pain suspicious to their clinician for SOD pain but normal ducts, no pancreatitis, and few with limited elevation in biochemistries, the placebo response was 37%; this might have been higher using the PGIC as part of EPISOD’s composite primary outcome.^3^ The observed outcomes are similar to: 1) the placebo response in a randomized trial of a protease inhibitor for painful chronic pancreatitis, and 2) the response to endoscopic therapy in an open-label randomized trial compared to surgery.^32, 33^ Other factors which may increase the placebo response include the patient’s expectation of how much the intervention will help and confidence with which the clinician explains the efficacy of their intervention.^28, 29, 34, 35^ We did not measure the white coat effect, but more than half of patients expected complete or near complete resolution of their symptoms. The magnitude of the intervention (ERCP and its known risks) and the patient’s high expectation imply a substantial placebo response. An optimist could reasonably argue that a helpful placebo is still helpful… It is reassuring that despite the rate of serious adverse events within 30 days of ERCP, very few patients (< 6%) believed they were worse 12 months later. Another placebo-controlled randomized trial would be necessary to differentiate the sphincterotomy response from the placebo response.

### Physician-defined characteristics of SOD correlate poorly with response

For decades, physicians have used duct dilation and biochemistries as objective markers for impaired drainage through the sphincter of Oddi. In contrast to historic trials of SOD,^5, 6^ the present study demonstrates that physician-defined characteristics of SOD correlate poorly with response to sphincterotomy and is consistent with prior observations.^36^ The reasons are undoubtedly multifactorial. First, the more widespread use of MRCP and endoscopic ultrasound means RESPOnD excluded patients with choledocholithiasis who might have been classified as SOD in older cohorts. Second, it is more common for gastroenterologists to consult on patients with abdominal pain who are already prescribed opioids for nonmalignant indications – abdominal pain or otherwise.^37^ Opioids are a major confounder since they cause bile duct dilation, elevated liver chemistries, and even acute pancreatitis; 44% of recent opioid users had a bile duct ≥ 12 mm, as compared to 31% for those without. Furthermore, recent opioids are more likely to associate with opioid-induced central sensitization, disruption to the gut microbiome, and their GI side effects overlap with the symptoms that mimic SOD; none of these are likely to help the chance of benefiting from sphincterotomy.

### The importance of pain characteristics in response

Nearly half of RESPOnD participants met criteria for underlying somatization at the time of enrollment. This likely represents a combination of individuals with a primary functional somatic syndrome and those who have developed central sensitization due to their chronic abdominal pain.^27^ Differentiating these pain subtypes is beyond the scope of this study, but the finding that 72% of individuals without somatization improved is important because a placebo response is unlikely to explain this completely. Similarly, patients least impacted by their pain, defined by normal mental and physical health, and who did not require opioids to manage their pain experienced response rates > 70%. These are populations who seem best to consider offering a high-risk intervention such as ERCP.

### Acute pancreatitis

Approximately half of RESPOnD participants presented with a history of acute pancreatitis prior to their index ERCP. While a natural history group is lacking in RESPOnD, the high (31%) recurrence rate of pancreatitis at a median of 6 months after ERCP suggests sphincterotomy is ineffective in reducing the rate of acute pancreatitis in this population. The observed recurrence rate is remarkably like an earlier randomized trial comparing biliary and pancreatic sphincterotomy for patients with iRAP^4^ and worse than the reported recurrence rate (15.1%) from a meta-analysis of 17 studies and 4,754 individuals classified as idiopathic acute pancreatitis.^38^ Having acute pancreatitis prior to the index ERCP was clearly associated with a higher likelihood of developing acute pancreatitis during follow-up and even comparable to patients whose pancreatitis is attributed to alcohol misuse and fail to quit.^39–42^ RESPOnD’s prospective study design with scheduled research follow-up assessments might have caused an inflation of the observed rate of AP when compared to studies whose approach to follow-up is more passive (e.g., a measurement bias). It is also noteworthy that 3% of individuals with no prior history of acute pancreatitis developed an attack during the first 12 months of follow-up. All of this is in addition to a significant risk of post-ERCP pancreatitis (10.3%). For patients with iRAP, these data argue against a short-term benefit of ameliorating subsequent pancreatitis episodes.

### Limitations

The principal strengths of RESPOnD include its multicenter and prospective study design with use of validated questionnaires to define baseline patient characteristics and PROMs. Although some patients were lost to follow-up before reaching the 12-month primary outcome, the success rate was similar in a sensitivity analysis restricted to patients with complete follow-up data. Additionally, the success rate did not change substantially when considering alternatives to the PGIC, including a reduction in pain-related disability using the same definition from the EPISOD trial (RAPID score of < 6). These observations reassure that the imputation method did not introduce bias and that the study’s primary outcome was reproducible using other PROMs. The use of neuromodulators and antispasmodics within 30 days of enrollment was low (49% and 28%, respectively). While this is likely due to these agents’ lack of efficacy when used earlier in their clinical course (thus prompting consultation at an endoscopy referral center), the overall rate of pharmacotherapy failure at the time of enrollment was not queried due to concerns about recall bias. Another limitation is the heterogeneity of the patient cohort, including some patients who did not have a history of cholecystectomy and a mixture of patients with biliary, pancreatic, or both subtypes of SOD. Although SOD is classically defined as a post-cholecystectomy syndrome, the study team elected to include these cases to maximize the study’s generalizability since the decision to perform ERCP was left to the discretion of the patient and treating physician based on the suspicion that their abdominal pain was originating in the biliary tree, pancreas, or both. Our subgroup analyses do not suggest that classically defined SOD subtypes perform differently. A history of cholecystectomy was not associated with a difference in success following the index ERCP (data not shown); 6 patients proceeded to cholecystectomy during follow-up, and all 4 who completed follow up at 12 months reported improvement. Similarly, patients with a history of acute pancreatitis as part of their SOD phenotype experienced similar response rates (by PROMs) to those with classically defined biliary SOD. Last, there is a real possibility of referral bias in this cohort. Patients referred to tertiary centers for suspected SOD may represent those with the most severe phenotype – and thus most susceptible to central sensitization and cross-sensitization. It is possible that there are unmeasured confounders which influenced the response rate. Despite its multicenter design and 4 years of recruitment, the final cohort was smaller than originally anticipated due to changes in practice patterns at participating centers and other unmeasured factors, raising the possibility of type II statistical error.

## Conclusion

RESPOnD demonstrates that nearly 60% patients who undergo ERCP for suspected SOD feel better after 12 months of follow-up, although the component attributable to a placebo response is unclear. Physician-defined characteristics such as duct size, lab tests, and an objective history of acute pancreatitis correlate poorly with a person’s likelihood of benefiting from the procedure, meaning the traditional Rome III classification system is a poor guide to deciding who should be offered ERCP. Despite high technical success rates of ERCP with sphincterotomy, patients with antecedent acute pancreatitis are still likely to develop a recurrent attack. The most important prognostic factors for response to ERCP are determined by the patient, more nuanced, and many are difficult to precisely diagnose in the clinic: those without underlying somatization – a surrogate measure of disordered pain processing and/or central sensitization – no recent use of opioids, and least disabled by their symptoms stand the greatest odds of improvement. Those who are least desperate for intervention seem to stand the best chance of improvement. RESPOnD highlights the importance of a careful history, the need for more objective pain biomarkers, and additional research into the placebo response of ERCP when performed for pain. In the meantime, clinicians should continue to tread cautiously and avoid the temptation to offer ERCP enthusiastically when “objective” features of SOD are present: they may open the door to a slippery slope.

## Supporting information

RESPOnD Study Protocol

## Data Availability

All data produced in the present study are available upon reasonable request to the authors

## Authorship

Gregory A Cote, MD, MS (Conceptualization: Lead; Formal analysis: Supporting;

Funding acquisition: Lead; Investigation: Lead; Methodology: Lead; Writing – original draft: Lead; Writing – review & editing: Equal)

B. Joseph Elmunzer (Investigation: Equal; Writing – review & editing: Equal) Haley Nitchie (Project administration: Lead)

Richard S. Kwon (Investigation: Equal; Writing – review & editing: Equal)

Field F. Willingham (Investigation: Equal; Writing – review & editing: Equal)

Sachin Wani (Investigation: Equal; Writing – review & editing: Equal) Vladimir Kushnir (Investigation: Equal; Writing – review & editing: Equal)

Amitabh Chak (Investigation: Equal; Writing – review & editing: Equal)

Vikesh Singh (Investigation: Equal; Writing – review & editing: Equal)

Georgios Papachristou (Investigation: Equal; Writing – review & editing: Equal)

Adam Slivka (Investigation: Equal; Writing – review & editing: Equal)

Shyam Varadarajulu (Investigation: Equal; Writing – review & editing: Equal)

Martin Freeman (Investigation: Equal; Writing – review & editing: Equal)

Srinivas Gaddam (Writing – review & editing: Equal)

Priya Jamidar (Writing – review & editing: Equal)

Paul Tarnasky (Investigation: Equal; Writing – review & editing: Equal)

Lydia D. Foster (Data curation: Lead; Formal analysis: Lead; Funding acquisition: Supporting; Writing – review & editing: Equal)

Peter B. Cotton (Conceptualization: Supporting; Formal analysis: Supporting; Writing – review & editing: Equal)

## Conflict of interest disclosure

No authors have a relevant conflict of interest.

**Guarantor of the article: Gregory Cote** is accepting full responsibility for the conduct of the study and has had access to the data and control of the decision to publish.

## Acronyms

SOD: Sphincter of Oddi disorders
DGBI: Disorders of gut-brain interaction
CBD: Common bile duct
iRAP: Idiopathic recurrent acute pancreatitis
MASLD: Metabolic dysfunction associated steatotic liver disease
RESPOnD: Results of Ercp for SPhincter of Oddi Disorders
PROM: Patient-reported outcome measures
SVI: Stent vs. Indomethacin trial
IBS: Irritable bowel syndrome
ULN: Upper limit of normal
PGIC: Patient Global Impression of Change
RAPID: Recurrent Abdominal Pain Intensity and Disability
EPISOD: Evaluating Predictors & Interventions in Sphincter of Oddi Dysfunction

## References

1. Cotton PB, Elta GH, Carter CR, et al. Rome IV. Gallbladder and Sphincter of Oddi Disorders. Gastroenterology 2016;150:1420–1429.

2. Hreinsson JP, Tornblom H, Tack J, et al. Factor Analysis of the Rome IV Criteria for Major Disorders of Gut-Brain Interaction (DGBI) Globally and Across Geographical, Sex, and Age Groups. Gastroenterology 2023;164:1211–1222.

3. Cotton PB, Durkalski V, Romagnuolo J, et al. Effect of endoscopic sphincterotomy for suspected sphincter of Oddi dysfunction on pain-related disability following cholecystectomy: the EPISOD randomized clinical trial. JAMA 2014;311:2101–9.

4. Cote GA, Imperiale TF, Schmidt SE, et al. Similar efficacies of biliary, with or without pancreatic, sphincterotomy in treatment of idiopathic recurrent acute pancreatitis. Gastroenterology 2012;143:1502–1509 e1.

5. Toouli J, Roberts-Thomson IC, Kellow J, et al. Manometry based randomised trial of endoscopic sphincterotomy for sphincter of Oddi dysfunction. Gut 2000;46:98–102.

6. Geenen JE, Hogan WJ, Dodds WJ, et al. The efficacy of endoscopic sphincterotomy after cholecystectomy in patients with sphincter-of-Oddi dysfunction. N Engl J Med 1989;320:82–7.

7. Behar J, Corazziari E, Guelrud M, et al. Functional gallbladder and sphincter of oddi disorders. Gastroenterology 2006;130:1498–509.

8. Guda NM, Trikudanathan G, Freeman ML. Idiopathic recurrent acute pancreatitis. Lancet Gastroenterol Hepatol 2018;3:720–728.

9. Barakat MT, Banerjee S. Incidental biliary dilation in the era of the opiate epidemic: High prevalence of biliary dilation in opiate users evaluated in the Emergency Department. World J Hepatol 2020;12:1289–1298.

10. Powell EE, Wong VW, Rinella M. Non-alcoholic fatty liver disease. Lancet 2021;397:2212–2224.

11. Ruhl CE, Everhart JE. Upper limits of normal for alanine aminotransferase activity in the United States population. Hepatology 2012;55:447–54.

12. Clark JM, Brancati FL, Diehl AM. The prevalence and etiology of elevated aminotransferase levels in the United States. Am J Gastroenterol 2003;98:960–7.

13. Ioannou GN, Boyko EJ, Lee SP. The prevalence and predictors of elevated serum aminotransferase activity in the United States in 1999-2002. Am J Gastroenterol 2006;101:76–82.

14. Elmunzer BJ, Foster LD, Serrano J, et al. Indomethacin with or without prophylactic pancreatic stent placement to prevent pancreatitis after ERCP: a randomised non-inferiority trial. Lancet 2024;403:450–458.

15. Suarez AL, Pauls Q, Durkalski-Mauldin V, Cotton PB. Sphincter of Oddi Manometry: Reproducibility of Measurements and Effect of Sphincterotomy in the EPISOD Study. J Neurogastroenterol Motil 2016;22:477–82.

16. Cote GA, Nitchie H, Elmunzer BJ, et al. Characteristics of Patients Undergoing Endoscopic Retrograde Cholangiopancreatography for Sphincter of Oddi Disorders. Clin Gastroenterol Hepatol 2022;20:e627–e634.

17. Durkalski V, Stewart W, MacDougall P, et al. Measuring episodic abdominal pain and disability in suspected sphincter of Oddi dysfunction. World J Gastroenterol 2010;16:4416–21.

18. Hays RD, Spritzer KL, Schalet BD, Cella D. PROMIS((R))-29 v2.0 profile physical and mental health summary scores. Qual Life Res 2018;27:1885–1891.

19. Banks PA, Bollen TL, Dervenis C, et al. Classification of acute pancreatitis--2012: revision of the Atlanta classification and definitions by international consensus. Gut 2013;62:102–11.

20. Li P, Stuart EA, Allison DB. Multiple Imputation: A Flexible Tool for Handling Missing Data. JAMA 2015;314:1966–7.

21. Smith ZL, Shah R, Elmunzer BJ, Chak A. The Next EPISOD: Trends in Utilization of Endoscopic Sphincterotomy for Sphincter of Oddi Dysfunction from 2010-2019. Clin Gastroenterol Hepatol 2022;20:e600–e609.

22. Watson RR, Klapman J, Komanduri S, et al. Wide disparities in attitudes and practices regarding Type II sphincter of Oddi dysfunction: a survey of expert U.S. endoscopists. Endosc Int Open 2016;4:E941–6.

23. Thiruvengadam NR, Saumoy M, Schaubel DE, et al. Rise In First-Time ERCP For Benign Indications >1 Year After Cholecystectomy Is Associated With Worse Outcomes. Clin Gastroenterol Hepatol 2024.

24. Doran FS. The sites to which pain is referred from the common bile-duct in man and its implication for the theory of referred pain. Br J Surg 1967;54:599–606.

25. Kingham JG, Dawson AM. Origin of chronic right upper quadrant pain. Gut 1985;26:783–8.

26. Chapman WP, Herrera R, Jones CM. A comparison of pain produced experimentally in lower esophagus, common bile duct, and upper small intestine with pain experienced by patients with diseases of biliary tract and pancreas. Surg Gynecol Obstet 1949;89:573–82.

27. Bourke JH, Langford RM, White PD. The common link between functional somatic syndromes may be central sensitisation. J Psychosom Res 2015;78:228–36.

28. Dettori JR, Norvell DC, Chapman JR. The Art of Surgery: The Strange World of the Placebo Response. Global Spine J 2019;9:680–683.

29. Doherty M, Dieppe P. The “placebo” response in osteoarthritis and its implications for clinical practice. Osteoarthritis Cartilage 2009;17:1255–62.

30. Gu AP, Gu CN, Ahmed AT, et al. Sham surgical procedures for pain intervention result in significant improvements in pain: systematic review and meta-analysis. J Clin Epidemiol 2017;83:18–23.

31. Jonas WB, Crawford C, Colloca L, et al. To what extent are surgery and invasive procedures effective beyond a placebo response? A systematic review with meta-analysis of randomised, sham controlled trials. BMJ Open 2015;5:e009655.

32. Hart PA, Osypchuk Y, Hovbakh I, et al. A Randomized Controlled Phase 2 Dose-Finding Trial to Evaluate the Efficacy and Safety of Camostat in the Treatment of Painful Chronic Pancreatitis: The TACTIC Study. Gastroenterology 2023.

33. Issa Y, Kempeneers MA, Bruno MJ, et al. Effect of Early Surgery vs Endoscopy-First Approach on Pain in Patients With Chronic Pancreatitis: The ESCAPE Randomized Clinical Trial. JAMA 2020;323:237–247.

34. Rossettini G, Campaci F, Bialosky J, et al. The Biology of Placebo and Nocebo Effects on Experimental and Chronic Pain: State of the Art. J Clin Med 2023;12.

35. Jones MP, Guthrie-Lyons L, Sato YA, Talley NJ. Factors Associated With Placebo Treatment Response in Functional Dyspepsia Clinical Trials. Am J Gastroenterol 2023;118:685–691.

36. Freeman ML, Gill M, Overby C, Cen YY. Predictors of outcomes after biliary and pancreatic sphincterotomy for sphincter of oddi dysfunction. J Clin Gastroenterol 2007;41:94–102.

37. Keyes KM, Rutherford C, Hamilton A, et al. What is the prevalence of and trend in opioid use disorder in the United States from 2010 to 2019? Using multiplier approaches to estimate prevalence for an unknown population size. Drug Alcohol Depend Rep 2022;3.

38. Hajibandeh S, Jurdon R, Heaton E, et al. The risk of recurrent pancreatitis after first episode of acute pancreatitis in relation to etiology and severity of disease: A systematic review, meta-analysis and meta-regression analysis. J Gastroenterol Hepatol 2023;38:1718–1733.

39. Lankisch PG, Breuer N, Bruns A, et al. Natural history of acute pancreatitis: a long-term population-based study. Am J Gastroenterol 2009;104:2797–805; quiz 2806.

40. Yadav D, O’Connell M, Papachristou GI. Natural history following the first attack of acute pancreatitis. Am J Gastroenterol 2012;107:1096–103.

41. Ahmed Ali U, Issa Y, Hagenaars JC, et al. Risk of Recurrent Pancreatitis and Progression to Chronic Pancreatitis After a First Episode of Acute Pancreatitis. Clin Gastroenterol Hepatol 2016;14:738–46.

42. Das R, Clarke B, Tang G, et al. Endoscopic sphincterotomy (ES) may not alter the natural history of idiopathic recurrent acute pancreatitis (IRAP). Pancreatology 2016;16:770–7.

