## Supplemental Materials for "Sphincterotomy for Biliary Sphincter of Oddi Disorder and idiopathic Acute Recurrent Pancreatitis: THE RESPOND LONGITUDINAL COHORT"

**Supplemental Table 1.** **Validated, self-reported instruments used in RESPOnD to assess comorbidity**

| **Baseline characteristic** | **Instrument** |
| --- | --- |
| Quality of life | **Patient-Reported Outcomes Measurement Information System® (PROMIS-29, v. 2.0)^1^**   - Assessment of anxiety, depression, fatigue, pain interference, physical function, sleep disturbance, ability to participate in social roles and activities, and a single pain intensity item - Low QOL: T score ≤ 40 |
| Psychiatric illness | **PROMIS-29** |
| Pain related disability | **RAPID**^2^   - Pain-related disability (90-day recall) instrument that was developed, validated, and used for the EPISOD study |
| Pain character | **PROMIS Pain Quality^3^ - Neuropathic and Nociceptive**   - Assessment of pain quality |
| Depression | **Brief Symptom Inventory (BSI-18)^4^**   - Assessment of 9 symptom scales: somatization, obsessive compulsive, interpersonal sensitivity, depression, anxiety, hostility, phobi anxiety, paranoid ideation, psychoticism. - High likelihood of depression: T score ≥ 65 |
| Anxiety | **PROMIS-29, BSI-18**   - High likelihood of anxiety: T score ≥ 65 |
| Functional GI disorder | **Rome Foundation Diagnostic Questionnaires^5^** |
| Somatization | **BSI-18**   - Assessment of psychological distress and somatization - High likelihood of somatization: T score ≥ 65 |
| Patient Expectation of Response | **QBS (Questions Before Sphincterotomy)**   - Quantitative measure of the degree to which a patient expects to improve after ERCP |

**Supplemental Table 2. Serious Adverse Events within 30 days of ERCP for suspected SOD**

| SAE Summary table | | | # Subjects | |
| --- | --- | --- | --- | --- |
|  | | # Events | Total | |
|  | | Total N | N | Percent |
| All Subjects |  | . | 213 | . |
| All Events |  | 109 | 68 | 31.9% |
| Blood and lymphatic system disorders | Anemia | 3 | 3 | 1.4% |
|  | Leukocytosis | 5 | 4 | 1.9% |
| Gastrointestinal disorders | Abdominal pain | 44 | 43 | 20.2% |
|  | Diarrhea | 1 | 1 | 0.5% |
|  | Duodenal perforation | 1 | 1 | 0.5% |
|  | Melena | 1 | 1 | 0.5% |
|  | Nausea | 4 | 4 | 1.9% |
|  | Pancreatic pseudocyst rupture | 1 | 1 | 0.5% |
|  | **Post-ERCP acute pancreatitis** | **24** | **22** | **10.3%** |
|  | Small intestinal obstruction | 1 | 1 | 0.5% |
| General disorders and administration site conditions | Generalized edema | 1 | 1 | 0.5% |
|  | Systemic inflammatory response syndrome | 1 | 1 | 0.5% |
| Hepatobiliary disorders | Bile duct obstruction | 1 | 1 | 0.5% |
| Injury, poisoning, and procedural complications | Burns second degree | 1 | 1 | 0.5% |
| Investigations | Lipase increased | 1 | 1 | 0.5% |
| Metabolism and nutrition disorders | Hypokalemia | 1 | 1 | 0.5% |
| Musculoskeletal and connective tissue disorders | Costochondritis | 1 | 1 | 0.5% |
| Neoplasms benign, malignant, and unspecified (incl cysts and polyps) | Adenocarcinoma pancreas | 1 | 1 | 0.5% |
| Nervous system disorders | Encephalopathy | 1 | 1 | 0.5% |
|  | Syncope | 1 | 1 | 0.5% |
| Psychiatric disorders | Confusional state | 2 | 2 | 0.9% |
| Renal and urinary disorders | Acute kidney injury | 3 | 2 | 0.9% |
| Respiratory, thoracic, and mediastinal disorders | Acute respiratory failure | 1 | 1 | 0.5% |
|  | Dyspnea | 1 | 1 | 0.5% |
|  | Hypoxia | 3 | 2 | 0.9% |
|  | Respiratory failure | 2 | 2 | 0.9% |
| Vascular disorders | Flushing | 1 | 1 | 0.5% |
|  | Hypotension | 1 | 1 | 0.5% |

**Supplemental Table 3. Correlation between primary outcome and secondary outcomes**

| **Secondary outcome** | **Overall** | | **Success by primary outcome** | | **Failure by primary outcome** | **Relative risk*** | **P value** |
| --- | --- | --- | --- | --- | --- | --- | --- |
| Change in RAPID score | -35.8 (-45.8, -25.7) | | -41.2  (-53.7, -28.7) | | -28.6  (-45.8, -11.4) | 0.76  (0.61, 0.94) | **0.0127** |
| **PROMIS-29** | | | | | | | |
| - Change in physical health summary score, mean (95% CI) | 4.7 (2.7, 6.6) | | 6.0 (3.8, 8.3) | | 2.8 (-0.4, 6.0) | 1.25 (1.02, 1.54) | **0.0294** |
| - Change in mental health summary score, mean (95% CI) | 5.7 (3.9, 7.6) | | 7.6 (5.2, 10.0) | | 3.2 (0.5, 5.9) | 1.30 (1.06, 1.61) | **0.0140** |
| **PROMIS-29 Sub-scores, mean (95% CI)**** | | | | | | | |
| - Physical function | 4.6 (2.7, 6.6) | | 6.1 (4.0, 8.3) | | 1.9 (-1.9, 5.8) | 1.34 (1.12, 1.62) | **0.0017** |
| - Anxiety | -2.4 (-4.4, -0.5) | | -3.5 (-5.9, -1.2) | | -0.4 (-3.8, 2.9) | 0.87 (0.74, 1.03) | 0.1090 |
| - Depression | 0.0 (-1.5, 1.6) | | -1.7 (-3.5, 0.0) | | 3.2 (0.6, 5.9) | 0.74 (0.60, 0.91) | **0.0038** |
| - Fatigue | -8.2 (-10.6, -5.9) | | -11.3 (-13.9, -8.7) | | -2.6 (-6.9, 1.6) | 0.76 (0.67, 0.86) | **<.0001** |
| - Sleep disturbance | -3.3 (-5.1,-1.5) | | -4.7 (-6.9, -2.6) | | -0.7 (-3.8, 2.4) | 0.81 (0.70, 0.93) | **0.0025** |
| - Ability to participate in social roles and activities | 7.3 (5.0,9.6) | | 8.6 (6.1, 11.0) | | 5.0 (0.4, 9.6) | 1.22 (1.04, 1.42) | **0.0139** |
| - Pain interference | -5.9 (-8.2, -3.7) | -7.9 (-10.5, -5.4) | | -2.3 (-6.5, 1.9) | 0.77 (0.67, 0.88) | 0.0002 | |

* per 30 point change for Change in RAPID score; per 10 point change for Change in PROMIS-29 score

**Unimputed; n=129 for physical function, anxiety and depression, n=130 for fatigue, sleep disturbance social roles, and pain interference; p-value indicates association with unimputed primary outcome.

**Supplemental Figure 1. Relationship between pain related disability and bile duct diameter**

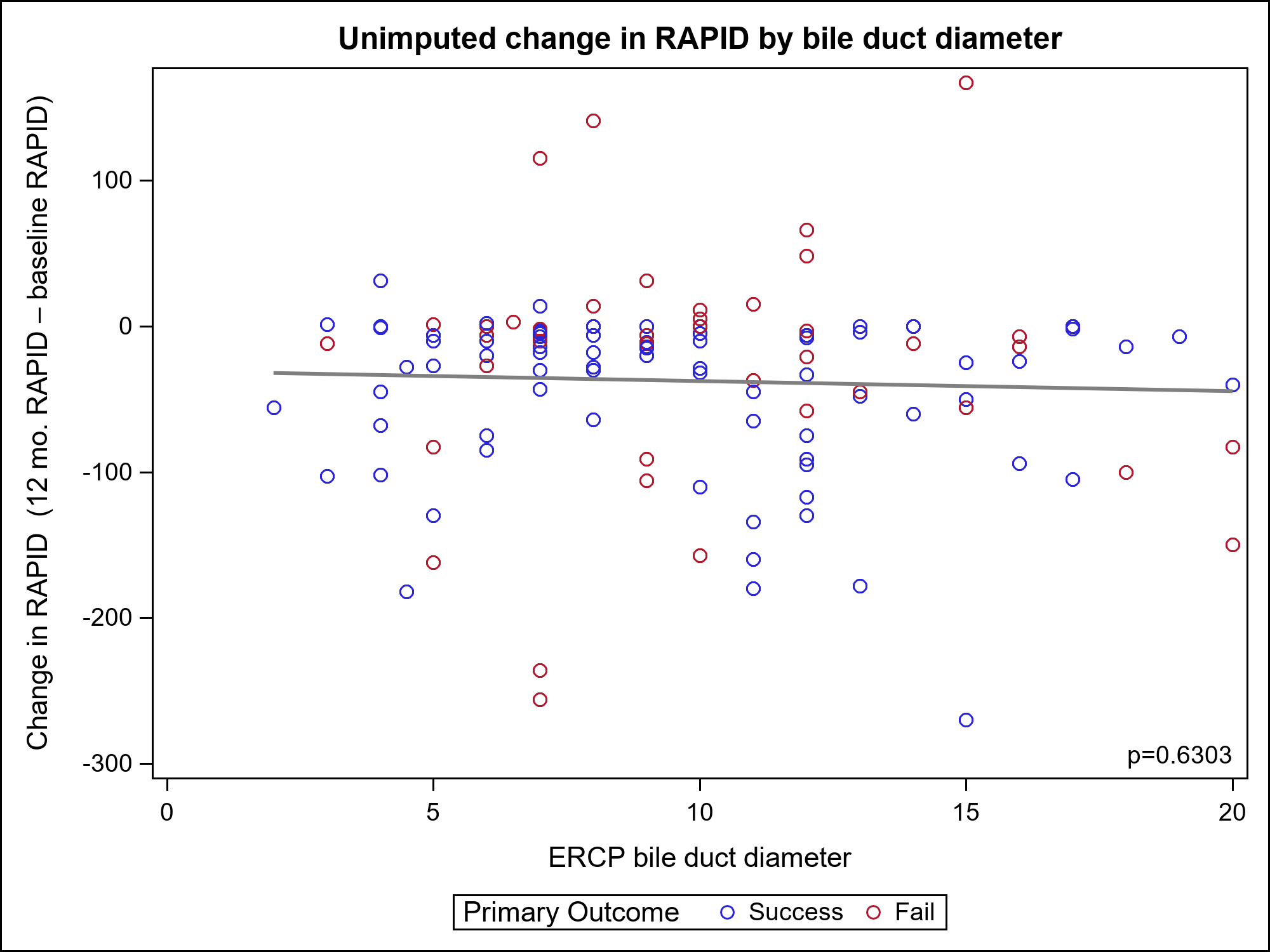

**Supplemental References**

1.	Hays RD, Spritzer KL, Schalet BD, Cella D. PROMIS((R))-29 v2.0 profile physical and mental health summary scores. Qual Life Res 2018;27:1885-1891.

2.	Durkalski V, Stewart W, MacDougall P, et al. Measuring episodic abdominal pain and disability in suspected sphincter of Oddi dysfunction. World J Gastroenterol 2010;16:4416-21.

3.	Askew RL, Cook KF, Keefe FJ, et al. A PROMIS Measure of Neuropathic Pain Quality. Value Health 2016;19:623-30.

4.	Derogatis LR, Melisaratos N. The Brief Symptom Inventory: an introductory report. Psychol Med 1983;13:595-605.

5.	Drossman DA, Hasler WL. Rome IV-Functional GI Disorders: Disorders of Gut-Brain Interaction. Gastroenterology 2016;150:1257-61.
