## Supplementary material for "Sphincterotomy for Biliary Sphincter of Oddi Disorder and idiopathic Acute Recurrent Pancreatitis: THE RESPOND LONGITUDINAL COHORT": RESPOnD Study Protocol

**Results of Ercp in SPhincter of Oddi Dysfunction:**

**The RESPOnD Study**

### Co-PRINCIPAL INVESTIGATORS

Gregory A. Cote

Peter B. Cotton

Lydia Foster

Protocol Version 2.0, 5-Mar-19

**SPONSOR**

National Institute of Diabetes and Digestive and Kidney Diseases (NIDDK)

**Division of Gastroenterology & Hepatology, Department of Medicine**

**and**

**The Data Coordination Unit (DCU)**

**Medical University of South Carolina**

**Charleston, SC 29425**

**Investigator's Agreement**

I have read the attached clinical protocol titled Results of Ercp in SPhincter of Oddi Dysfunction: The RESPOnD Study and dated March 5, 2019 and agree to conduct the protocol as written in this document.

I agree to comply with the Declaration of Helsinki/Tokyo/Venice on Experimentation in Humans as required by the United States Food and Drug Administration regulations; the Code of Federal Regulations Title 21 parts 50, 56, 312; the Code of Federal Regulations Title 45 part 46; ICH Good Clinical Practice Guidelines; and all other applicable guidelines.

I understand this document contains confidential information of the National Institute of Diabetes & Digestive and Kidney Diseases and The Data Coordination Unit and cannot be disclosed to anyone other than members of my staff conducting this trial and members of my Institutional Review Board or Ethical Committee.

I agree to ensure that this information will not be used for any purpose other than the evaluation or conduct of this clinical trial without the prior written permission of the National Institute of Diabetes & Digestive and Kidney Diseases and The Data Coordination Unit.

_________________________________	_____________________________

Signature of Principal Investigator	Date

_________________________________

Printed name of Principal Investigator

_________________________________	______________________________

Signature of Co-Principal Investigator	Date

(When applicable)

_________________________________

Printed name of Co-Principal Investigator

(When applicable)

1.	SUMMARY	1

2.	OBJECTIVES	1

2.1	Primary.	1

2.2	Secondary.	1

2.3	Exploratory.	1

3.	BACKGROUND AND RATIONALE	2

3.1	Background of Disease	2

3.2	Rationale	3

4.	STUDY PLAN	3

4.1	Study Design	3

4.2	Study Population	3

4.3	Study Sites	4

4.4	Estimated Study Duration	4

5.	ELIGIBILITY CRITERIA	4

5.1	Inclusion Criteria	4

5.2	Exclusion Criteria	4

6.	PARTICIPANT RECRUITMENT	4

6.1	Methods	4

7.	PARTICIPANT ENROLLMENT	5

7.1	Eligibility Assessment	5

7.2	Presentation of Informed Consent	6

8.	STUDY PROCEDURES	6

8.1	Baseline Assessments	6

8.1.1	Medical History and Record Review	6

8.1.2	Baseline assessment instruments	6

8.2	Treatment Procedures	7

8.2.1	ERCP procedure	7

8.2.2	Concomitant or Ancillary Therapy	8

8.3	Follow-up Procedure	8

8.3.1	30-day follow-up	8

8.3.2	Post-SVI follow-up	8

9.	DISCONTINUATION OF PARTICIPATION	9

9.1	Participant Withdrawal	9

9.2	Participant Removal from Study	9

9.3	Procedure for Discontinuation	10

9.4	Participant Transfers	10

10.	OUTCOMES DEFINITIONS	10

10.1	Primary	10

10.2	Secondary	10

10.2.1	Change in RAPID score from baseline	10

10.2.2	Change in PROMIS 29 Profile from baseline	10

10.3	Exploratory	10

11.	DATA MANAGEMENT	11

11.1	Site Monitoring	11

11.2	Data Management	11

11.3	Data Security and Confidentiality	11

11.4	Data Quality Assurance	12

12.	STATISTICAL CONSIDERATIONS	12

12.1	Sample Size and Power Estimation	12

12.2	Statistical Analysis	12

13.	REGULATORY AND ETHICAL OBLIGATIONS	13

13.1	Informed Consent	13

13.2	Institutional Review Board (IRB)	14

13.2.1	Initial Review and Approval	14

13.2.2	Amendments	14

13.2.3	Annual	14

14.	ADMINISTRATIVE AND LEGAL OBLIGATIONS	14

14.1	Study Termination	14

14.2	Study Documentation and Storage	15

14.3	Publication Policy	15

15.	STUDY ORGANIZATION	15

15.1	Steering Committee	15

15.2	Executive Committee	16

15.3	Data Safety and Monitoring Committee	16

16.	REFERENCES	17

17.	Study related definitions	18

17.1	Acute pancreatitis.	18

17.2	Recurrent acute pancreatitis (RAP).	18

17.3	Idiopathic RAP.	18

17.4	Calcific chronic pancreatitis.	18

17.5	Functional Sphincter of Oddi Disorder.	18

18.	APPENDICES	18

18.1	List of Abbreviations	18

1. SUMMARY

This is a longitudinal cohort study of patients undergoing endoscopic retrograde cholangiopancreatography (ERCP) for suspected sphincter of Oddi disorders. We estimate that approximately 360 patients will be enrolled into this cohort study, and be followed for 12 months after ERCP to assess for their response to the procedure. The study period will be 5 years.

2. OBJECTIVES
   2.1 Primary.

To measure the benefit of ERCP in patients with Functional Sphincter of Oddi Disorders.

The primary outcome is measuring the patient-reported benefit of ERCP (level of improvement since the procedure) using the 12-month Patient Global Impression of Change. This metric will be compared to additional patient-centered outcomes, using validated instruments for pain, self-perceived quality of life, and pain-related disability.

2.2 Secondary.

To determine factors associated with response to endoscopic sphincterotomy.

Functional Sphincter of Oddi disorders are defined by the presence of symptoms with one or more objective abnormalities: common bile duct dilation, transiently abnormal liver chemistries with pain, or idiopathic acute pancreatitis (historically type I or II Sphincter of Oddi Dysfunction (SOD)). The impact of these factors on response to ERCP is based on older and limited prospective data. Furthermore, there are several other covariates likely to influence the probability of response.

We hypothesize that the objective findings currently used to define Functional Sphincter of Oddi Disorders can be enhanced by the development of comprehensive predictive models that include other probable covariates. Among others, these may include smoking status, symptom duration, opiate utilization, pain characteristics, the severity and pattern of pain-related disability, presence of concurrent functional GI symptoms, and indication for and response to previous cholecystectomy. To test this hypothesis, we will define the relationship between current factors (lab abnormalities, duct diameter, and prior acute pancreatitis) and response to ERCP, as defined by aim #1. We will then develop one or more models incorporating other covariates to better define factors associated with response to ERCP. This may result in a weighted scale that could be used to guide clinicians on who are most likely to benefit from ERCP.

2.3 Exploratory.

Among patients with Functional Pancreatic Sphincter of Oddi Disorder, measure the acute pancreatitis incidence rate ratio.

The incidence rate of acute pancreatitis is variable, and a probable predictor of having recurrent attacks after ERCP. Among patients with idiopathic recurrent acute pancreatitis, a goal of ERCP is to reduce the risk of subsequent acute pancreatitis bouts by: 1) identifying occult etiologies for acute pancreatitis and, 2) performing sphincterotomy when no other obstructive lesion is found. Prospective data are limited.

In the subgroup of enrollees with Functional Pancreatic Sphincter of Oddi Disorder (i.e., idiopathic recurrent acute pancreatitis), we will measure the incidence rate ratio of acute pancreatitis (episodes/time prior to ERCP ÷ episodes/time following ERCP). These point estimates will be useful for future studies evaluating ERCP and other interventions in this population.

3. BACKGROUND AND RATIONALE
   3.1 Background of Disease

The concept that SOD can cause pain and attacks of acute pancreatitis by raising bile and pancreatic duct intraductal pressure is intuitive and has been a fervent topic of debate for nearly a century.^1, 2^ Each year in the U.S., thousands of patients are assigned a diagnosis of “suspected SOD.” SOD has been considered in patients with persistent or recurrent biliary pains after undergoing cholecystectomy, and also in patients with idiopathic acute pancreatitis. Cholecystectomy is a common operation: at least 700,000 are performed each year in the U.S., and the number has risen progressively since the introduction of the laparoscopic method. More than 10% of the patients present later with “post-cholecystectomy pain,” many of whom are subjected to ERCP with biliary, pancreatic, or dual sphincterotomy to treat SOD.^3^ In this setting, the benefit of ERCP and sphincterotomy is unpredictable, and the risks are substantial. ERCP procedures cause acute pancreatitis in 10-15% of cases, even in expert hands and when appropriate prophylactic measures are utilized.^4^

The clinical approach to patients with suspected SOD has been greatly influenced by the early work of the Milwaukee group, who promoted the use of ERCP with sphincter of Oddi manometry and manometry-directed sphincterotomy. They proposed a categorization of biliary SOD almost 30 years ago. Type I patients had definite evidence for biliary obstruction (dilated bile duct and abnormal liver enzymes). Type II had only one of these criteria, and Type III had neither. These categories were embraced by the clinical community and by the Rome foundation, an international organization which attempts to refine the diagnosis and management of functional digestive disorders every 5-10 years. The 3rd iteration of the Rome consensus was published in 2006, and included the type I, II, and III classification.^5^

More recently, the concept that SOD can cause biliary pain in patients without any objective abnormalities (a.k.a., type III SOD) was strongly contested by the “Evaluating Predictors & Interventions in Sphincter of Oddi Dysfunction” (EPISOD) study, which irrefutably demonstrated no difference in outcomes between patients who underwent sphincterotomy or sham treatment at ERCP; furthermore, there was no prognostic value of sphincter of Oddi manometry, meaning that the finding of elevated basal sphincter pressure had no association with baseline pain characteristics and no impact on the likelihood of response to sphincterotomy.^6, 7^ SOD has been implicated as a “cause” for idiopathic recurrent acute pancreatitis (also known as pancreatic SOD). The clinical significance of pancreatic SOD as a cause for unexplained acute pancreatitis, and the therapeutic role of pancreatic sphincterotomy was challenged by an 89-patient, open label, randomized trial of ERCP with sphincterotomy for patients with idiopathic recurrent acute pancreatitis.^8^

3.2 Rationale

Due in large part to the results of EPISOD, the Rome IV consensus changed the criteria for functional sphincter of Oddi disorders and eliminated the concept of type III SOD.^3^ It also recommended that type II biliary SOD be designated Functional Biliary Sphincter Disorder (FBSD) (table 1). The Rome IV criteria still rely heavily on abnormal blood chemistries and post-cholecystectomy bile duct diameter. These are the only “objective criteria” used to distinguish patients who might still benefit from ERCP from those who clearly do not (those historically defined as type III SOD). A similar condition, functional pancreatic sphincter of Oddi disorder (FPSD), is considered for patients with idiopathic, recurrent acute pancreatitis.

| **Table 1. Rome IV definitions for Functional Sphincter of Oddi Disorders^3^** | |
| --- | --- |
| **Diagnostic criteria for Functional Biliary Sphincter of Oddi disorder (FBSD)** | **Diagnostic criteria for Functional Pancreatic Sphincter of Oddi Disorder (FPSD)** |
| 1. Criteria for biliary pain, as defined by Rome IV 2. Elevated liver chemistries or dilated bile duct, but not both 3. Absence of bile duct stones or other structural abnormalities | 1. Documented recurrent episodes of acute pancreatitis 2. Other etiologies of pancreatitis excluded 3. Negative endoscopic ultrasound 4. Abnormal sphincter of Oddi manometry |

We suspect that publication of the EPISOD study results has had a major impact on clinical practice, but many patients with FBSD or FPSD are still being referred to tertiary centers. Those meeting an updated Rome IV definition continue to undergo ERCP and sphincterotomy despite limited data supporting its benefit. The outcomes following ERCP for this indication are unclear, and which patients to treat remain controversial. Evidence that the clinical community is confused about this was confirmed by recent surveys.^9-11^

4. STUDY PLAN
   4.1 Study Design

The Results of Ercp in SPhincter of Oddi Dysfunction (RESPOnD) study will be a stringent, longitudinal cohort study of patients undergoing ERCP for “suspected SOD.” Our overarching hypothesis is that responders to sphincterotomy can be defined more precisely than the current Rome IV definition for Functional Sphincter of Oddi Disorders.

4.2 Study Population

Patients who have suspected SOD or idiopathic, recurrent acute pancreatitis will be considered for participation in the RESPOnD study.

4.3 Study Sites

All sites participating in the SVI trial will recruit patients for the RESPOnD study. To bolster recruitment, non-SVI sites will be incorporated into the RESPOnD consortium and recruit patients meeting the RESPOnD eligibility criteria.

4.4 Estimated Study Duration

The study duration will be five years.

5. ELIGIBILITY CRITERIA
   5.1 Inclusion Criteria
(1) Adult patients (ages 18 and over) or their authorized legally acceptable representative must consent to be in the study and must have signed and dated an approved consent form, which conforms to federal and institutional guidelines.
(2) *Only for sites participating in the SVI trial:* Relationship to SVI trial (either of the following):
a. Screened to participate in the SVI trial prior to ERCP
b. Randomized in the SVI trial within the past 12 months
(3) Indication for ERCP procedure (either or both of the following):
a. Suspected sphincter of Oddi dysfunction per treating physician
b. Idiopathic, recurrent acute pancreatitis per RESPOnD definition
   5.2 Exclusion Criteria
(1) Prior endoscopic sphincterotomy before enrollment
(2) No plan to undergo ERCP
(3) Known chronic calcific pancreatitis
(4) Known main pancreatic duct stricture
(5) Known main pancreatic duct stone
(6) Pregnant or breast-feeding at the time of proposed study entry
(7) Psychiatric disorders that would preclude obtaining informed consent
6. PARTICIPANT RECRUITMENT
   6.1 Methods

The methods used for recruitment of participants in the study will be devoid of any procedures that may be construed as coercive. The recruitment process will not involve any restrictions on sociodemographic factors including age, gender, or ethnic characteristics of the participant population. However, the composition of the study participant population will depend on patient referral sources available to the Clinical Centers.

Women and minority populations will be included, consistent with NIH guidelines (<https://grants.nih.gov/grants/funding/women_min/women_min.htm>).

Participants will be recruited through the gastrointestinal clinical practices of each of the participating Clinical Centers. After standard evaluation in the clinics, patients who have pain, and who may be categorized as suffering from SOD or unexplained recurrent pancreatitis will be interviewed by the PI or a research coordinator to establish eligibility criteria and obtain consent. Consent will be obtained in a private location after the patient has had time to ask any questions.

7. PARTICIPANT ENROLLMENT
   7.1 Eligibility Assessment

All patients screened to participate in the SVI trial, or subjects who have been randomized in the SVI trial within the past 12 months will be screened for eligibility for entry into the RESPOnD study. For sites not participating in the SVI trial, all patients referred for ERCP with the indication of “suspected SOD” or “idiopathic recurrent acute pancreatitis” should be screened to participate in RESPOnD.

As in all trials, the goal is to achieve a high level of compliance with protocol requirements by assuring, during the eligibility assessment, that the potential participant is fully informed and agrees to the protocol requirements. With the exception of SVI subjects who have already been randomized at the inception of the RESPOnD study, every effort must be undertaken to enroll subjects prior to their SVI ERCP procedure, as applicable. In addition, participants with a strong likelihood of non-adherence due to inability to complete the self-administered forms or comply with the planned follow-up procedures, should not knowingly be registered. Adherence of the clinical center staff to careful assessment of the participant’s understanding of the study and a clinical center environment which supports the continued commitment of the participants are essential for the study to be successfully completed.

Pre-randomization or pre-registration evaluation of all patients will include:

(1) Verification that all inclusion/exclusion criteria listed in Section 5 have been evaluated correctly.
(2) Completion of all applicable pre-study assessments.
(3) Evaluation and documentation of relevant medical and surgical history including indication for, timing of, and response to cholecystectomy.
(4) Completion of physical examination and documentation of the required data.
(5) Verification that all required information has been documented, and copies of all pertinent reports (e.g. prior liver and pancreas chemistries, bile and pancreatic duct characteristics on prior cross-sectional imaging, and prior cholecystectomy findings) have been obtained.
(6) Obtaining signed and dated informed consent.
    7.2 Presentation of Informed Consent

Consent will be obtained by either the Principal Investigator or by individuals approved by the Clinical Center's Principal Investigator. The initial consent should be the IRB-approved version corresponding to the version of the protocol approved when the screening was initiated. Informed consent is to be obtained from the participant in a private location. Participants will be given time to read the consent document and ask questions.

8. STUDY PROCEDURES
   8.1 Baseline Assessments
       8.1.1 Medical History and Record Review

Study-relevant medical history will be reviewed and documented. This will include questions about past medical history, including the indication for ERCP, indication for and response to prior cholecystectomy, and details regarding prior pancreatitis episodes (if applicable).

8.1.2 Baseline assessment instruments

Subjects will complete a series of instruments designed to characterize their pain, pain-related disability, and presence of potential confounding factors such as underlying somatization, depression, anxiety, patient expectation of response, coexisting functional disorders, and opiate utilization (table 2). In total, we estimate the baseline visit will require approximately one hour of face-to-face time with the patient.

| **Table 2. Baseline assessment instruments for RESPOnD study** | | | |
| --- | --- | --- | --- |
| **Potential covariate(s)** | **Instrument(s)** | **Explanation** | **Expected time to complete* (min)** |
| Patient characteristics | RESPOnD study case report forms | - Data will be collected as applicable and available: prior gallstone disease, liver/pancreas chemistries, duct characteristics on prior imaging, body mass index, waist circumference, presence of hepatic steatosis, other prior abdominal surgery, personal history of fibromyalgia | 5 (with patient)  60 (coordinator time, including abstract of prior records) |
| Functional syndrome assessment | Rome Foundation Diagnostic Questionnaires ^12^ | - Irritable bowel syndrome (IBS) - Biliary pain and Sphincter of Oddi disorders | 10 |
| Current pain disability | RAPID^13^ | - RAPID: Pain-related disability (90-day recall); developed, validated, and used for the EPISOD study | 3 |
| Pain character | PROMIS Pain Quality - Neuropathic and Nociceptive | - Assessment of pain quality | 2-5 |
| Quality of life | PROMIS 29 Profile | - Assessment of anxiety, depression, fatigue, pain interference, physical function, sleep disturbance, ability to participate in social roles and activities, and a single pain intensity item | 3 |
| Somatization | Brief Symptom Inventory - 18 | - Assessment of psychological distress and somatization | 5 |
| Patient expectation of response | QBS (Questions Before Sphincterotomy) | - Quantitative measure of the degree to which a patient expects to improve after ERCP | 1 |
| Medication utilization | Ancillary case report forms | - Use of opiates and other specified medications in the past 30 days - Number of days using opiates and other specified medications in the past 30 days | 5 |
| * Estimated time for a patient, working with a dedicated RESPOnD study coordinator, to complete the document(s) (in minutes).  SVI = Stent vs. Indomethacin; RAPID = Recurrent Abdominal Pain Intensity and Disability  **The study investigator or their staff will respond to any subjects presenting suicidal ideations by using standard psychiatric procedures for safety purposes. If the BSI -18 Question 17 is answered low (1-2), the Coordinator will refer the subject to a mental health specialist for follow up. If the risk is considered moderate to high (3-4), the Coordinator will page the PI or a clinician and seek guidance for possible admission to mental health services without leaving the subject alone. | | | |

8.2 Treatment Procedures
    8.2.1 ERCP procedure

During ERCP, the physician will perform all diagnostic and therapeutic maneuvers per clinical judgement. Diagnostic maneuvers may include: 1) cholangiogram, 2) pancreatogram, 3) sphincter of Oddi manometry, or some combination. Relevant data on ERCP diagnostics will be collected, and include diameters of the common bile duct and pancreatic duct, presence and characteristics of relevant pathology such as stricture or stone, and basal sphincter of Oddi pressure by manometry in the biliary and pancreatic sphincter segments, if performed.

Therapeutic maneuvers may include: 1) biliary sphincterotomy, 2) pancreatic sphincterotomy, 3) common bile duct stent placement, 4) pancreatic duct stent placement, 5) orifice dilation, 6) stricture dilation, 7) stone extraction, or some combination. Relevant data will be collected regarding sphincterotomy technique, stent characteristics (diameter, length, and composite material), dilation technique and maximal diameter, and stone location and size, as applicable.

8.2.2 Concomitant or Ancillary Therapy

Throughout the study, any concomitant medications or treatments necessary to provide adequate supportive care may be prescribed.

8.3 Follow-up Procedure

The goal of the study is to achieve complete, accurate follow-up until completion of the trial. Appropriate compliance strategies will be implemented at clinical centers to encourage and support participants in protocol adherence. The ultimate success of this trial will depend upon the timely submission of complete and accurate data on all follow-up forms.

8.3.1 30-day follow-up
      8.3.1.1 SVI subjects

No additional follow-up specific to the RESPOnD study, as post-procedure events are captured within the existing SVI framework.

8.3.1.2 Subjects not enrolled in SVI

Subjects will be contacted by the local RESPOnD research coordinator 30±5 days after enrollment or their index ERCP procedure (whichever occurs later) and assessed for serious adverse events.

8.3.2 Post-30 day follow-up

Subjects will be contacted by a research coordinator centralized at the Clinical Coordinating Center at 3, 6, 9, and 12 months after enrollment. During each of these encounters, several instruments will be used to assess response to the index ERCP *(table 3)*. Subjects will be compensated $50 for their time completing the questionnaires at months 6 and 12. Serious Adverse Events related to progress of the presenting conditions will be collected on a separate CRF.

| **Table 3. RESPOnD study follow-up assessment instruments** | | | | | | |
| --- | --- | --- | --- | --- | --- | --- |
| **Instrument(s)** | **Explanation** | **Follow-up visit (month)** | | | | **Expected time to complete (min)** |
|  |  | **3** | **6** | **9** | **12** |  |
| Patient Global Impression of Change (PGIC) | Patient’s assessment of efficacy from the index ERCP procedure | x | x | x | x | 1 |
| RAPID | Pain-related disability (90-day recall), which includes a pain intensity score | x | x | x | x | 3 |
| PROMIS 29 Profile | Overall assessment of physical and mental health |  | x |  | x | 3 |
| PROMIS Pain Quality - Neuropathic and Nociceptive | Qualitative assessment of pain |  |  |  | x | 2-5 |
| RESPOnD Follow-up case report form | Health care evaluations for further pancreatobiliary interventions  Pancreatitis events* | x | x | x | x | 5 |
| Medication utilization | - Use of opiates and other specified medications in the past 30 days - Number of days using opiates and other specified medications in the past 30 days - Initiation and response to opiates and other specified medications for their GI symptoms | x | x | x | x | 5 |
| * Medical records relevant to pancreatitis events will be reviewed by a site investigator, to validate that each episode meets the RESPOnD definitions outlined in section 17 of this protocol. | | | | | | |

9. DISCONTINUATION OF PARTICIPATION
   9.1 Participant Withdrawal

The participant has the right to voluntarily withdraw from the study at any time for any reason without prejudice to his/her future medical care by the physician or at the institution.

For the occasional participant who withdraws consent, the date and reason for consent withdrawal should be documented. Participant data will be included in the analysis up to the date of the consent withdrawal.

A distinction should be made between participants who fail to complete all forms on schedule or who miss some clinic visits and the withdrawal of consent. Missed or rescheduled visits will be documented, but the participant will continue to be followed in the future according to protocol requirements, and all follow-up data will be included in the analysis.

9.2 Participant Removal from Study

Participants may be removed from the study if any one or more of the following events occur:

(1) Significant protocol violation or noncompliance, either on the part of the participant or Investigator.
(2) Refusal of the participant to continue treatment and/or observations.
(3) If the physician or sponsor believes it is in the participant's best interest to discontinue participation in the study.
(4) Administrative reasons, e.g., sponsor termination of the study.

Non-adherence to the protocol intervention will be appropriately documented, but will not be a reason for discontinuing the participant in the protocol.

9.3 Procedure for Discontinuation

The procedure to be followed at the time a participant either discontinues participation or is removed from the study is:

(1) Check for the development of adverse events.
(2) Complete the End-of-Study form and include an explanation of why the participant is withdrawing or withdrawn.
(3) Attempt to perform follow-up evaluations if indicated.
    9.4 Participant Transfers

Whenever a participant's medical care transfers to another clinical setting, every attempt must be made to obtain continued follow-up data and information on self-administered forms. DCU should be notified when this occurs, so that appropriate arrangements can be made, wherever possible for the participant to continue to participate in the study.

10. OUTCOMES DEFINITIONS
    10.1 Primary

PGIC (Patient Global Impression of Change) “much improved” or “very much improved” at month 12, with no repeat ERCP or surgical procedure on the sphincter of Oddi, biliary tree, or pancreas during the follow-up period and who has the same or less days of prescription analgesic use during month 12 compared to baseline.

10.2 Secondary
     10.2.1 Change in RAPID score from baseline
     10.2.2 Change in PROMIS 29 Profile from baseline
10.3 Exploratory

This outcome is restricted to subjects with idiopathic, recurrent acute pancreatitis at the time of enrollment.

Incidence rate ratio, defined as the number of acute pancreatitis episodes/months prior to ERCP ÷ number of acute pancreatitis episodes/months following ERCP

11. DATA MANAGEMENT
    11.1 Site Monitoring

Study data will be monitored on a routine basis for completeness, timeliness, logic, and consistency by utilizing internal quality controls (e.g., built in range and logic checks in WebDCU™), external quality controls (via statistical programming), and data clarification queries. In addition, each subject’s signed informed consent form and HIPAA Authorization Form will be remotely reviewed.

11.2 Data Management

Data management will be handled by the Data Coordination Unit (DCU) in the Department of Public Health Sciences at the Medical University of South Carolina (MUSC). All study activities will be conducted in coordination with the study PI, the clinical sites, and NIH, and will use an electronic data acquisition method where all clinical data on randomized subjects will be entered by the site personnel in real time. The latest version of each CRF will be available as a PDF file on the study website for use as worksheets and source documents by study personnel.

The study data will be managed (including data queries) by the DCU using the WebDCU™ system. This user-friendly web-based database system, developed by the DCU, will be used for regulatory document management, data entry, data validation, project progress monitoring, subject tracking, site monitoring, user customizable report generation and secure data transfer. Upon entry of CRFs into the study database, quality control procedures will be applied at each stage of data handling in order to ensure compliance with GCP guidelines, integrity of the study data, and document processing system reliability.

11.3 Data Security and Confidentiality

During the course of the study, user access to the files with subject identifiers, and files with study outcomes will be restricted to core staff with any exceptions to be approved by the Executive Committee.

In addition to use of passwords and other security measures, all documents containing identifying information on individuals or physicians are considered confidential materials and will be safeguarded to the greatest possible extent. No information, which identifies a specific person, hospital, or physician, will be released to, or discussed with anyone other than study staff members.

Because the DCU uses a web-based system, source documents and CRFs will remain at the participating sites. The study database only identifies study subjects by unique study identification codes. All data will be stored in a manner that is HIPAA compliant, without the ability to track the information back to a specific subject except through a password protected system. All collected information about a subject will be stored by a unique identification code. All DCU personnel are certified by the NIH Office of Human Subjects Research in the Protection of Human Research Subjects course.

12. STATISTICAL CONSIDERATIONS

**Refer to the RESPOnD Statistical Analysis Plan for additional details.**

12.1 Sample Size and Power Estimation

Sample size estimation for the proposed study is based on a combination of projected enrollment in the parent trial (SVI), anticipated success rates, and the primary outcome measure (see section 10.1). Based on the SVI trial, there is the following information: 1) The maximum original sample size of the SVI trial is 1430; 2) The current rate of non-redo SOD subjects who are enrolled in SVI, thereby meeting this ancillary study’s enrollment criteria, is 33%; 3) The projected remaining SVI enrollment at the anticipated start date for the proposed study (January 2018) is 814-879, with 240-300 enrolled in the prior 12 months; 5% of subjects who are eligible for RESPOnD will refuse to participate in SVI but consider enrolling into RESPOnD. Based on this information we anticipate that there will be approximately 385 subjects eligible to be enrolled in the proposed study. Taking into account a small percentage that will refuse consent to participate in a long-term observational study, we estimate that 360 subjects will be enrolled into this study.

12.2 Statistical Analysis

The unadjusted primary outcome success proportion (see section 10.1) will be estimated with a two-sided 95% confidence interval. The secondary patient-reported outcome measures (change in RAPID and PROMIS 29 Profile scores from baseline) will be treated as continuous variables. The correlation between each outcome and the primary outcome status (success/failure) will be examined to assess the strength of association. The correlation will be examined over time using a mixed effects model and fitted using SAS PROC MIXED.

Pre-defined potential predictors of the primary (dichotomous) outcome will be tested for univariate association. Univariate tests with a p-value ≤0.10 will prompt inclusion of these variables in a multivariable logistic model to determine if there are specific prognostic variables of good outcome (success) that can be further explored in future studies or included in a weighted scale to guide clinicians.

In the subset of patients with FPSD who meet criteria for idiopathic recurrent acute pancreatitis at baseline, the incidence will be estimated as number of acute pancreatitis episodes/time following ERCP and will be compared to the incidence prior to ERCP. We will also examine the time to first episode.

13. REGULATORY AND ETHICAL OBLIGATIONS
    13.1 Informed Consent

In accordance with US Federal and ICH Good Clinical Practice Consolidated Guideline, it is the investigator’s responsibility to ensure that informed consent is obtained from the participant or participant’s legally authorized representative before participating in an investigational study, after an adequate explanation of the purpose, methods, risks, potential benefits and participant responsibilities of the study. Procedures that are to be performed as part of the practice of medicine and which would be done whether or not study entry was contemplated, such as for diagnosis or treatment of a disease or medical condition, may be performed and the results subsequently used for determining study eligibility without first obtaining consent. On the other hand, informed consent must be obtained prior to initiation of any screening procedures that are performed solely for the purpose of determining eligibility for research.

The original signed consent must be retained in the institution’s records and is subject to review by the sponsor, DCU, the FDA or representative from another agency that performs the same function, and the IRB responsible for the conduct of the institution. All elements listed in the ICH Good Clinical Practice guidelines must be included in the informed consent.

Informed consent will be obtained by either the Principal Investigator or by individuals approved by the Clinical Center’s Principal Investigator and whose names have been submitted to DCU. Informed consent will be obtained from the participant or participant’s legally acceptable representative after the details of the protocol have been reviewed. The individual responsible for obtaining consent will assure, prior to signing of the informed consent, that the participant has had all questions regarding therapy and the protocol answered.

13.2 Institutional Review Board (IRB)

In accordance with US federal regulations and ICH Good Clinical Practice Consolidated Guideline) all research involving human subjects and changes to the research plan must be reviewed and approved by an IRB.

13.2.1 Initial Review and Approval

A copy of the protocol, proposed informed consent form, other written participant information, and any proposed advertising material must be submitted to the Clinical Center’s IRB for written approval. A copy of the IRB approval of the protocol and informed consent form must be received by DCU before recruitment of participants into the study.

13.2.2 Amendments

Protocol amendments and changes to the informed consent template may only be made with the prior approval of the RESPOnD Executive Committee, the NIDDK, and the SVI DSMB. The Site Principal Investigators must agree to all protocol amendments and revisions to the informed consent document as dictated by the RESPOnD Executive Committee and approved by the SVI DSMB and NIDDK. The RESPOnD leadership team will submit amendments to the IRB for approval. Evidence of DSMB and NIDDK approval of the amendment will be included as part of the submission to the IRB.

13.2.3 Annual

The Principal Investigator will be responsible for obtaining annual IRB approval renewal throughout the duration of the study.

14. ADMINISTRATIVE AND LEGAL OBLIGATIONS
    14.1 Study Termination

The study will be complete when all patients have had their final study assessments. The Sponsor or Executive Committee reserves the right to terminate the study if new information becomes available on the safety or efficacy of the study product or if such action is justified.

If the study is terminated, the investigator will provide any outstanding data or documentation (e.g., case report form pages) considered appropriate by DCU at the time.

The Clinical Center reserves the right to terminate the study according to the contract. The investigator is responsible for notifying the IRB in writing of the trial’s completion or early termination. A copy of the notification must be sent to DCU.

14.2 Study Documentation and Storage

Source documents are the original or valid records of participant information from which case report form data are obtained. These include, but are not limited to, reports of test results, hospital charts and medical records, diaries, and correspondence. Case report form entries may be considered source data if the case report form is the site of original notation, such as the patient satisfaction questionnaire or quality of life instrument.

The study requires the Principal Investigator at each Clinical Center to retain a comprehensive and centralized filing system of all study related documentation, suitable for inspection by DCU, FDA representatives or a representative from another agency which performs the same function. Study documentation should be maintained after study termination according to the appropriate time frame as mandated by federal regulations. No study document should be destroyed without prior written agreement between the Principal Investigator and DCU. Should the Principal Investigator wish to assign the study records to another party or move them to another location, written agreement must be obtained from DCU.

14.3 Publication Policy

Investigators will be offered the opportunity to publish as a group or with recognition of individual authors. This decision will be made before analyses are conducted. Refer to the study Publication Policy for more details.

15. STUDY ORGANIZATION
    15.1 Steering Committee

The RESPOnD Steering Committee will be comprised of the RESPOnD Executive Committee (see section 15.2 for details), all RESPOnD Clinical Center Principal Investigators, and Dr. Doug Drossman who is serving as a consultant to the RESPOnD study. Peter Cotton will serve as Chair of the RESPOnD Steering Committee. The Steering Committee will communicate no less than once monthly to discuss progress on recruitment and retention; webinars may be conducted quarterly or more frequently as needed, to discuss barriers toward recruitment and retention.

15.2 Executive Committee

The RESPOnD Executive Committee will be comprised of the RESPOnD Principal Investigators (Cote, Cotton and Foster), the SVI Study Chair (Joe Elmunzer), the SVI lead biostatistician (Valerie Durkalski-Mauldin), one rotating RESPOnD Clinical Center Principal Investigator, and one or more representatives from the NIDDK as designated). The Executive Committee will communicate no less than once quarterly throughout the study.

15.3 Data Safety and Monitoring Committee

The monitoring of data quality and subject safety in this trial will be overseen by the existing SVI Data and Safety Monitoring Board (DSMB). The SVI DSMB members are appointed by the NIH.

The members will have a meeting with the PI and study statistician prior to study commencement to discuss the protocol as well as content and format of DSMB reports. The DCU will prepare the requested reports at the pre-specified time intervals. Both open and closed reports will be distributed – open reports will be available to the Executive Committee members and will be blinded to treatment assignment while closed reports will only be available to the DSMB members and will only be unblinded upon request by the DSMB members.

16. REFERENCES

1.	Weir JF, Snell AM. Symptoms that persist after cholecystectomy: their nature and probable significance. JAMA 1935;105:1093-1098.

2.	Ivy AC, Sandblom P. Biliary dyskinesia. Ann Intern Med 1934;8:115-122.

3.	Cotton PB, Elta GH, Carter CR, et al. Rome IV. Gallbladder and Sphincter of Oddi Disorders. Gastroenterology 2016;150:1420-1429.

4.	Dumonceau JM, Andriulli A, Elmunzer BJ, et al. Prophylaxis of post-ERCP pancreatitis: European Society of Gastrointestinal Endoscopy (ESGE) Guideline - updated June 2014. Endoscopy 2014;46:799-815.

5.	Behar J, Corazziari E, Guelrud M, et al. Functional gallbladder and sphincter of oddi disorders. Gastroenterology 2006;130:1498-509.

6.	Cotton PB, Durkalski V, Romagnuolo J, et al. Effect of endoscopic sphincterotomy for suspected sphincter of Oddi dysfunction on pain-related disability following cholecystectomy: the EPISOD randomized clinical trial. JAMA 2014;311:2101-9.

7.	Romagnuolo J, Cotton PB, Durkalski V, et al. Can patient and pain characteristics predict manometric sphincter of Oddi dysfunction in patients with clinically suspected sphincter of Oddi dysfunction? Gastrointest Endosc 2014;79:765-72.

8.	Cote GA, Imperiale TF, Schmidt SE, et al. Similar efficacies of biliary, with or without pancreatic, sphincterotomy in treatment of idiopathic recurrent acute pancreatitis. Gastroenterology 2012;143:1502-1509 e1.

9.	Watson RR, Klapman J, Komanduri S, et al. Wide disparities in attitudes and practices regarding Type II sphincter of Oddi dysfunction: a survey of expert U.S. endoscopists. Endosc Int Open 2016;4:E941-6.

10.	Suarez AL, Cotton PB, Pauls Q, Elmunzer BJ, Durkalski-Mauldin V, Cote GA. Sphincter of Oddi dysfunction: A survey of current practice in USA. Gastroenterology & Hepatology International Journal 2016;1.

11.	Suarez AL, Kutlu O, Cunningham SC, Bingener J, Morgan KA, Cotton PB. How Much Pain Relief Do Patients Expect After Cholecystectomy? Abstract. Gastroenterology 2016;150:S1257.

12.	Drossman DA, Hasler WL. Rome IV-Functional GI Disorders: Disorders of Gut-Brain Interaction. Gastroenterology 2016;150:1257-61.

13.	Durkalski V, Stewart W, MacDougall P, et al. Measuring episodic abdominal pain and disability in suspected sphincter of Oddi dysfunction. World J Gastroenterol 2010;16:4416-21.

17. Study related definitions
    17.1 Acute pancreatitis.

The definition of acute pancreatitis will be per consensus (Atlanta guidelines):(Banks, Bollen et al. 2013) “The diagnosis of acute pancreatitis requires two of the following three features: (1) abdominal pain consistent with acute pancreatitis (acute onset of a persistent, severe, epigastric pain often radiating to the back); (2) serum lipase activity (or amylase activity) at least three times greater than the upper limit of normal; and (3) characteristic findings of acute pancreatitis on CECT and less commonly MRI or transabdominal ultrasonography.”

17.2 Recurrent acute pancreatitis (RAP).

RAP will be defined as two or more discrete episodes of acute pancreatitis that occur >30 days apart with complete recovery from the first before commencement of the second episode.

17.3 Idiopathic RAP.

A patient with RAP will be defined as idiopathic if no etiology is evident to the treating physician after a thorough history, physical examination, routine laboratories (serum calcium, lipids (triglyceride level < 500mg/dL), liver chemistries), and cross-sectional imaging (transabdominal ultrasound and/or CECT). Patients with a history of current or previous smoking will be considered idiopathic if a second risk factor is absent.

17.4 Calcific chronic pancreatitis.

This is defined as parenchymal or ductal calcifications identified on computed tomography or magnetic resonance imaging scan.

17.5 Functional Sphincter of Oddi Disorder.

The definition of a Functional Sphincter of Oddi Disorder will be based on the Rome IV criteria detailed in table 1 of this protocol.

18. APPENDICES
    18.1 List of Abbreviations

ERCP	Endoscopic Retrograde CholangioPancreatography

SVI	Stent vs. Indomethacin Trial

SOD	Sphincter of Oddi Dysfunction

EPISOD	Evaluating Predictors & Interventions in Sphincter of Oddi Dysfunction

FBSD	Functional Biliary Sphincter Disorder

FPSD	Functional Pancreatic Sphincter Disorder

RESPOnD	Results of Ercp in SPhincter of Oddi Dysfunction

DCU	Data Coordination Unit

IBS	Irritable Bowel Syndrome

RAPID	Recurrent Abdominal Pain Intensity and Disability

QBS	Questions Before Sphincterotomy

PGIC	Patient Global Impression of Change

CRFs	Case Report Forms

MUSC	Medical University of South Carolina
